# Predictors of Response and Rational Combinations for the Novel MCL-1 Inhibitor MIK665 in Acute Myeloid Leukemia

**DOI:** 10.1101/2024.11.07.24316814

**Authors:** Joseph Saad, Rhiannon Newman, Elmira Khabusheva, Sofia Aakko, Eric Durand, Mahesh Tambe, Heikki Kuusanmäki, Alun Parsons, Juho J. Miettinen, Komal Kumar Javarappa, Nemo Ikonen, Mika Kontro, Kimmo Porkka, Heiko Maacke, Janghee Woo, Ensar Halilovic, Caroline A. Heckman

**Affiliations:** Institute for Molecular Medicine Finland (FIMM), Helsinki Institute of Life Science (HiLIFE), iCAN Digital Precision Cancer Medicine Flagship, University of Helsinki, Helsinki, Finland; Novartis Biomedical Research, Basel, Switzerland; Department of Hematology, Helsinki University Hospital Comprehensive Cancer Center, Helsinki, Finland; Foundation for the Finnish Cancer Institute, Helsinki, Finland; Winship Cancer Institute of Emory University, Atlanta, GA, USA; Novartis Biomedical Research, Cambridge, MA, USA

**Author notes:** RN and EK contributed equally to this study. **Correspondence:** Caroline Heckman, PhD, Institute for Molecular Medicine Finland (FIMM) University of Helsinki, P.O. Box 20 (Tukholmankatu 8), 00014 Helsinki, Finland. **Competing interests:** Sofia Aakko is currently an employee of Faron Pharmaceuticals, and Eric Durand is an employee of Owkin. During their participation in this study, Eric Durand, Janghee Woo, Heiko Maacke, and Ensar Halilovic were employees and shareholders of Novartis. Caroline Heckman received research funding from Novartis related to this work and has received unrelated research funding from BMS/Celgene, Kronos Bio, Oncopeptides, Orion Pharma, WntResearch, and the Innovative Medicines Initiative 2 project HARMONY, plus personal fees from Amgen and Autolus. Mika Kontro has received personal fees from Astellas Pharma, AbbVie, Bristol-Myers Squibb, Faron Pharmaceuticals, Novartis and Pfizer outside the submitted work. Kimmo Porkka has received unrelated research funding from Incyte, Novartis and Roche.

## Abstract

Despite promising anti-leukemic activity of MCL-1 inhibitors in preclinical studies of acute myeloid leukemia (AML), their progress through clinical evaluation has in part been challenged by limited knowledge of patient subgroups suitable for treatment. To stratify patients with AML for MCL-1 inhibitor-based treatment, we evaluated the sensitivity of 42 primary AML samples to MCL-1 inhibitor MIK665 (S64315) and contrasted their molecular profiles. We observed that MIK665 sensitive samples had a more differentiated phenotype, whereas resistant samples displayed higher levels of ABCB1 (MDR1) and the anti-apoptotic protein BCL-XL. Further evaluation revealed that *ABCB1* expression has good predictive performance in identifying MIK665 primary resistant samples. To induce sensitivity, we treated MIK665-resistant samples with ABCB1 inhibitor elacridar, BCL-XL inhibitor A1331852, or BCL-2 inhibitor venetoclax in combination with MIK665. While combinations with elacridar and A1331852 were not effective, the combination of MIK665 and venetoclax effectively eliminated AML blasts compared to either of the agents alone. Additionally, the combination restored sensitivity of samples with primary venetoclax resistance. Overall, this study indicates that elevated *ABCB1* expression is a potential predictor of resistance to MIK665 in AML, and that a combination of MIK665 with venetoclax may be effective for overcoming resistance to either MCL-1 or BCL-2 inhibition.

## Introduction

Of the several novel targeted therapies recently approved for the treatment of acute myeloid leukemia (AML), the combinations of azacitidine or low dose cytarabine with the BCL-2 inhibitor venetoclax for newly diagnosed patients unfit for intensive chemotherapy are particularly efficacious for this previously difficult to treat subgroup of patients (1–4). Apoptosis is a tightly regulated molecular process essential for the maintenance of cellular homeostasis, and the deregulation of which is a fundamental hallmark of tumorigenesis (5). The dependency of AML blasts on different anti-apoptotic BCL-2 family members, such as BCL-2, MCL-1, and BCL-XL, to escape apoptosis mark these proteins as ideal targets for therapeutic inhibition (6). Despite significant improvements in overall survival and remission rates, venetoclax-based therapies are challenged by both primary and acquired resistance leading to relapse (7). This raises the need for novel, rational drug combinations capable of attaining deeper and prolonged responses.

In AML, elevated MCL-1 expression plays a central role in escape from apoptosis and has been associated with poor patient prognosis (8,9). As such, inhibitors targeting MCL-1 have been evaluated in preclinical studies alone and in combination with BCL-2 inhibitors resulting in encouraging findings (10–13). These results have supported the clinical evaluation of several MCL-1 inhibitors as monotherapies or in combination with BCL-2 inhibitors and hypomethylating agents (14,15). However, the progression of MCL-1 inhibitors towards clinical approval has been hindered by dose-limiting cardiotoxicity and limited knowledge of biomarkers of response (14). Therefore, identification of patient subgroups likely to benefit from MCL-1 inhibition, as well as suitable drug combinations, could improve efficacy and accelerate the development of this drug class.

In this study, we evaluated the activity of the MCL-1 inhibitor MIK665 in AML preclinical models. To identify indicators of response, we assessed the *ex vivo* sensitivity of 42 AML samples to MIK665 and compared the transcriptional and protein expression profiles of MIK665 sensitive and resistant samples. The drug response groups were clearly distinguished by disease maturation phenotype and specific gene expression patterns. Additionally, we found that *ABCB1* expression is a predictive indicator of MIK665 resistance in AML samples. We also identified the combination of MIK665 and venetoclax as an effective strategy to overcome resistance to either agent in AML cell lines and samples. This study uncovers novel response patterns to MIK665 in AML that could aid the identification of patients that would benefit from MCL-1 inhibitor-based therapy and improve outcome.

## Materials and Methods

### Patient samples

Bone marrow (BM) aspirates and matched skin biopsies from patients with AML (n=42), as well as healthy BM control samples (n=2), were collected from the Helsinki University Hospital or the Finnish Hematology Registry and Biobank following written informed consent, according to protocols approved by the local ethics committee (permit numbers 239/13/03/00/2010 and 303/13/03/01/2011), and in compliance with the Declaration of Helsinki. BM mononuclear cells (MNCs) were isolated by Ficoll density gradient centrifugation (GE Healthcare, Buckinghamshire, UK) and suspended in conditioned medium (CM: RPMI 1640, 12.5% HS-5 conditioned medium, 10% fetal bovine serum (FBS), 2 mM L-glutamine, 100 units/ml penicillin and 100 µg/ml streptomycin) (16). All AML samples contained at least 50% malignant MNCs. RNA from the BM MNCs was prepared using the AllPrep DNA/RNA/miRNA Universal kit or miRNeasy mini kit (Qiagen, Hilden, Germany), and then subjected to RNA sequencing as previously described (17). Remaining MNCs from the samples were viably cryopreserved for further experiments. The clinical characteristics of the AML patient samples used are summarized in Table 1.

**Table 1.** Clinical characteristics of the FIMM AML patient cohort.

|  |  | **Total (n=42)** |
| --- | --- | --- |
| Median age (range), years |  | 63 (22-79) |
| Disease Stage | Diagnosis, n (%) | 36 (85.7%) |
|  | Relapse, n (%) | 4 (9.5%) |
|  | Refractory, n (%) | 2 (4.8%) |
| Median blast percentage (range) |  | 70.5 (50-95) |
| FAB | AML M0/1, n (%) | 14 (33.3%) |
|  | AML M2, n (%) | 8 (19%) |
|  | AML M4, n (%) | 3 (7.1%) |
|  | AML M5, n (%) | 5 (11.9%) |
|  | NA, n (%) | 12 (28.6%) |

FAB, French-American-British classification; NA, data not available

### Multiparamteric flow cytometry-based drug testing of patient samples

Viably cryopreserved AML and healthy MNCs were thawed, treated with DNase I, and suspended in CM for *ex vivo* drug testing. All samples were tested with MCL-1 inhibitors MIK665 and S63845, and BCL-2 inhibitor venetoclax. The drugs were dissolved in DMSO and pre-plated on 96-well V-bottom plates (Thermo Fisher Scientific, Carlsbad, CA) in 7 increasing concentrations from 0.1 to 1000 nM using an Echo 550 acoustic liquid handler (Labcyte, San Jose, CA, USA). Cells were incubated on the drug plates for 48 h at 37 °C in 5% CO_2_. Following initial profiling, selected AML samples (n=9) were taken forward for drug combination testing of MIK665 combined with venetoclax, ABCB1 inhibitor elacridar, or BCL-XL inhibitor A1331852. Each drug was tested in 6 increasing concentrations from 1 and 1000 nM, while the other drug was fixed at 30 nM (Supplementary Figure 1). When the number of cells from a patient sample was insufficient, the combination of MIK665 in increasing concentrations with the other drug fixed at 30 nM was prioritized. Following incubation with the drugs, cells were stained with an antibody panel designed for the identification of myeloid cell populations (Supplementary Table 1). Afterwards, the plates were analyzed using flow cytometry on the iQue Screener PLUS (Intellicyt, Albuquerque, NM, USA) and gating was performed using the ForeCyt software (Intellicyt). All drugs used in single agent and combination screens are summarized in Supplementary Table 2. The gating strategy used is illustrated in Supplementary Figure 2.

### Cell line expression and dependency analysis

Publicly available data was downloaded from DepMap portal and analyzed in GraphPad Prism. Gene expression data was available for 45 myeloid leukemia cell lines, and dependency data for 24 myeloid leukemia cell lines. A low Chronos score corresponds to a high gene dependency. Visualization of *BCL2*, *BCL2L1*, and *MCL1* expression and dependency was made for 24 cell lines which had both data sets available.

### Cell line culture

HEL, HL-60, MOLM-13, and Kasumi-1 cell lines were purchased from DSMZ (Braunschweig, Germany). MV4-11 cell line was purchased from ATCC (Manassas, VA, USA). HEL, Kasumi-1 and MV4-11 were maintained in RPMI 1640 medium supplemented with 10-20% heat-inactivated FBS, L-glutamine (2 mM), penicillin (100 U/mL), and streptomycin (100 mg/mL). All cells were maintained at 37 °C in 5% CO_2_.

### Generation of venetoclax-resistant (VenR) cell lines

Kasumi-1, MV4-11, MOLM-13, and HL-60 cells were exposed to increasing concentrations of venetoclax (from 12.5 nM – 1000 nM) with the drug concentration being doubled every 2 days, as previously described (18). VenR cell lines were derived from parental cells that continued to proliferate in the presence of 1000 nM of venetoclax.

### Knockout of *ABCB1* in HEL cells using CRISPR/Cas9

The gRNA targeting exon ENSE00003398270 of *ABCB1* (5’ – AAGTCCAGCCCCATGGATGA– 3’) was inserted into the lentiCRISPRv2GFP vector following the protocol as previously described by the Zhang lab (19). Lentiviral particles were generated by transfection of second-generation lentiviral systems into HEK293-FT cells by calcium phosphate transfection (Promega, Madison, WI, USA). The viral suspension was used to transduce HEL cells for 5 days at 37 °C and 5% CO_2_. Following this, fluorescence-activated cell sorting of GFP-positive cells into 96-well plates was carried out using the BD Influx cell sorter (Franklin Lakes, NJ, USA). Cells were maintained in 100 µl of complete RPMI media per well and expanded when needed. PCR was used to identify *ABCB1* knockout, which was confirmed by sanger sequencing and western blotting.

### CellTiterGlo-based drug testing of cell lines

Cell lines were tested with MIK665 and venetoclax as single agents or in combination in a range of 5 increasing concentrations from 6.25 to 100 nM (Supplementary Figure 3). The compounds were added to a 384-well plate using an acoustic liquid handling device Echo 550 (Labcyte). All cell lines were incubated on the drug plates at 37 °C and 5% CO_2_ for 48 h. Cell viability was measured by adding 25 µl of CellTiter-Glo reagent (Promega) to each well and luminescence signal read using a PHERAstar plate reader (BMG Labtech, Ortenberg, Germany).

### Data Analysis

For drug response assessment, viability readouts from drug treated wells were normalized to negative (DMSO) and positive (benzathonium chloride) controls, and inhibition dose-response curves were generated for each sample and treatment. Consequently, the modified integration of these curves yielded a drug sensitivity score (DSS) as previously described (20). The DSS is directly proportional to the response of a sample to a drug and has a range of 0 to 50. For drug combination synergy readouts, SynergyFinder 2.0 was used for the calculation of ZIP synergy scores (21).

Bulk RNA sequencing for AML patient samples was analyzed in-house as previously described, yielding a gene-sample raw count matrix (17,22). Trimmed Mean of M values (TMM) normalization was performed using the edgeR package and counts per million (CPM) values were computed (23). Differential gene expression analysis was performed using a limma-voom pipeline for regression modeling, identifying differentially expressed genes (FDR < 0.1) (24).

Confirmatory data analyses were performed using publicly available AML patient sample datasets: TCGA and BEAT AML (25,26).

All statistical tests were performed using R software (R version 4.0.0), and nonparametric tests used in cases where the normality of distribution by the Shapiro-Wilk test was not verified. Two group-comparisons were done by the 2-sample t-test or by the Mann Whitney U-test. Correlations between two continuous sets of values were performed using the Pearson or the Spearman method. Multiple testing correction by the Benjamini-Hochberg method was performed when applicable.

### Data Availability Statement

RNA sequencing data of AML samples is available in Supplementary Table 3, and drug sensitivity testing results from the single agent screen is available in Supplementary Table 6. For other original data, please contact the corresponding author.

For more detailed descriptions of the methodology, see the Supplementary Materials and Methods section. For the full list of western blot antibodies and RT-qPCR primers used, see Supplementary Tables 4 and 5.

## Results

### Primary AML cells display variable response to MIK665

Dose response of AML samples (n=42) to MCL-1 inhibitor MIK665 was assessed using a multiparametric flow cytometry-based assay. The leukocyte cell viability dose-response curve of each sample was converted into a drug sensitivity score (DSS), directly proportional to the sample’s response to MIK665 (Figure 1A). AML leukocyte populations demonstrated a DSS continuum across the samples, ranging from 5.2 to 34.9. Based on the DSS distribution, we selected DSS cutoffs of 10 and 20 to represent resistant (n=10) and sensitive (n=15) samples, respectively. The remaining 17 samples were defined as intermediate responders (Figure 1B). The effect of MIK665 on the leukocytes of a sensitive and a resistant AML sample is illustrated in Supplementary Figure 4. In parallel, S63845, an MCL-1 inhibitor structurally related to MIK665, was also tested in the samples resulting in a similar response pattern to MIK665 (Supplementary Figure 5) (27). The full list of results obtained from the single agent screening experiments is available in Supplementary Table 6.

**Figure 1.**
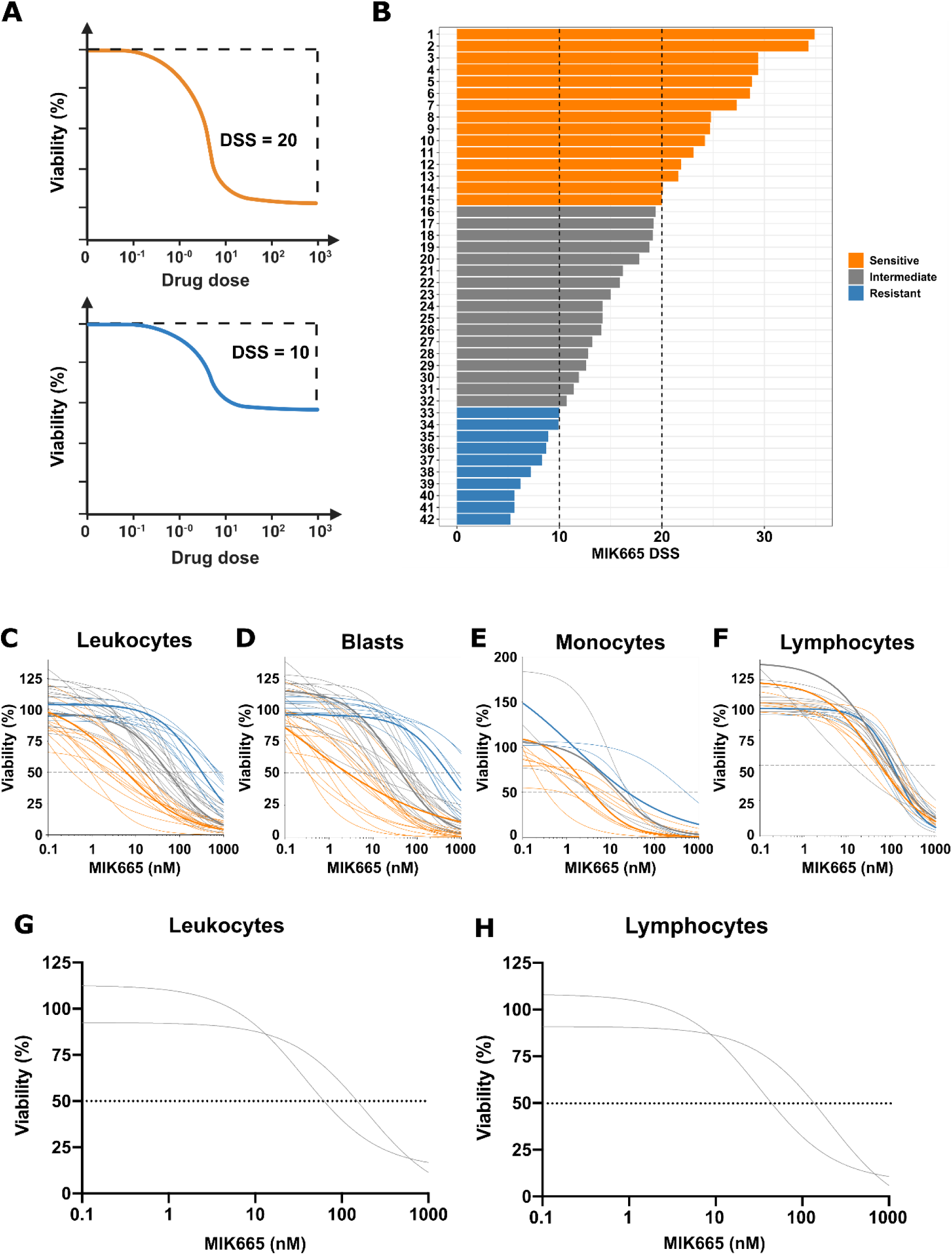
Response of AML bone marrow cell populations to MCL-1 inhibitor MIK665. **A)** An illustration showing the relationship between leukocyte cell viability dose response curves and the drug sensitivity score (DSS). The DSS is a modified area over the cell viability curve, which is directly proportional to the drug sensitivity of a sample. Created with BioRender.com. **B)** Waterfall plot showing the MIK665 DSS values in leukocytes of AML samples (n=42) in decreasing order. DSS cutoffs were used to create sample subgroups based on response to MIK665, such that DSS ≤ 10 was considered resistant (blue), DSS ≥ 20 was considered sensitive (orange), and 10 < DSS < 20 was considered intermediate (grey). MIK665 dose-response curves for **C)** leukocytes (relative IC50 values: 5.4 nM, 45 nM, and 134.8 nM for sensitive, intermediate, and resistant samples, respectively), **D)** blast cells (relative IC50 values: 2 nM, 44.1 nM and 158.4 nM respectively), **E)** monocytes (IC50 values: 3.8 nM, 14.5 nM, and 22.9 nM respectively), and **F)** lymphocytes (IC50 values: 52.6 nM, 97.3 nM, and 84.5 nM respectively). Representative samples with the median DSS from each group are highlighted by thicker lines on the dose-response curves. All cell populations were identified using flow cytometry. MIK665 activity was evaluated in 2 healthy BM samples, resulting in dose-response curves for **G)** leukocytes (DSS: 10 and 12, IC50: 192 nM and 36.4 nM) and **H)** lymphocytes (DSS: 10.7 and 14.2, IC50: 204.5 nM and 32.1 nM).

Viability dose-response curves for leukocyte (CD45+), blast (CD45dim-sidescatter low), monocyte (CD14+) and lymphocyte (CD45high-sidescatter low) subpopulations are illustrated in Figures 1C-F. Of the 42 AML samples, 16 contained a quantifiable monocytic population (representing ≥ 5% of the total leukocytes), 75% (n=12) of which were sensitive to MIK665 (Figure 1E). 23 out of the 42 samples contained a quantifiable lymphocyte population, 96% (n=22) of which were not sensitive to the treatment (Figure 1F). Additionally, MIK665 was tested on BM MNCs from 2 healthy donors. Intermediate DSS values (10 and 12) were observed for the leukocyte populations, with the lymphocyte populations showing similar responses, further indicating that MIK665 has a limited effect on healthy hematopoietic cells (Figure 1G-H). Together, these data indicated that MIK665 was particularly effective in AML cells differentiated towards the monocytic lineage, while its effect on lymphocytes was limited.

### High expression of ABCB1 or BCL-XL is associated with resistance to MIK665

To identify molecular indicators distinguishing MIK665-sensitive (n=15) and resistant (n=10) samples, we performed differential gene expression analysis. The genes analyzed were restricted to protein-coding genes having sufficiently high expression level (Supplementary Materials and Methods). 112 genes were differentially expressed (FDR < 0.1), 74 of which were upregulated and 38 were downregulated in MIK665-resistant compared to sensitive samples (Figure 2A, Supplementary Table 7).

**Figure 2.**
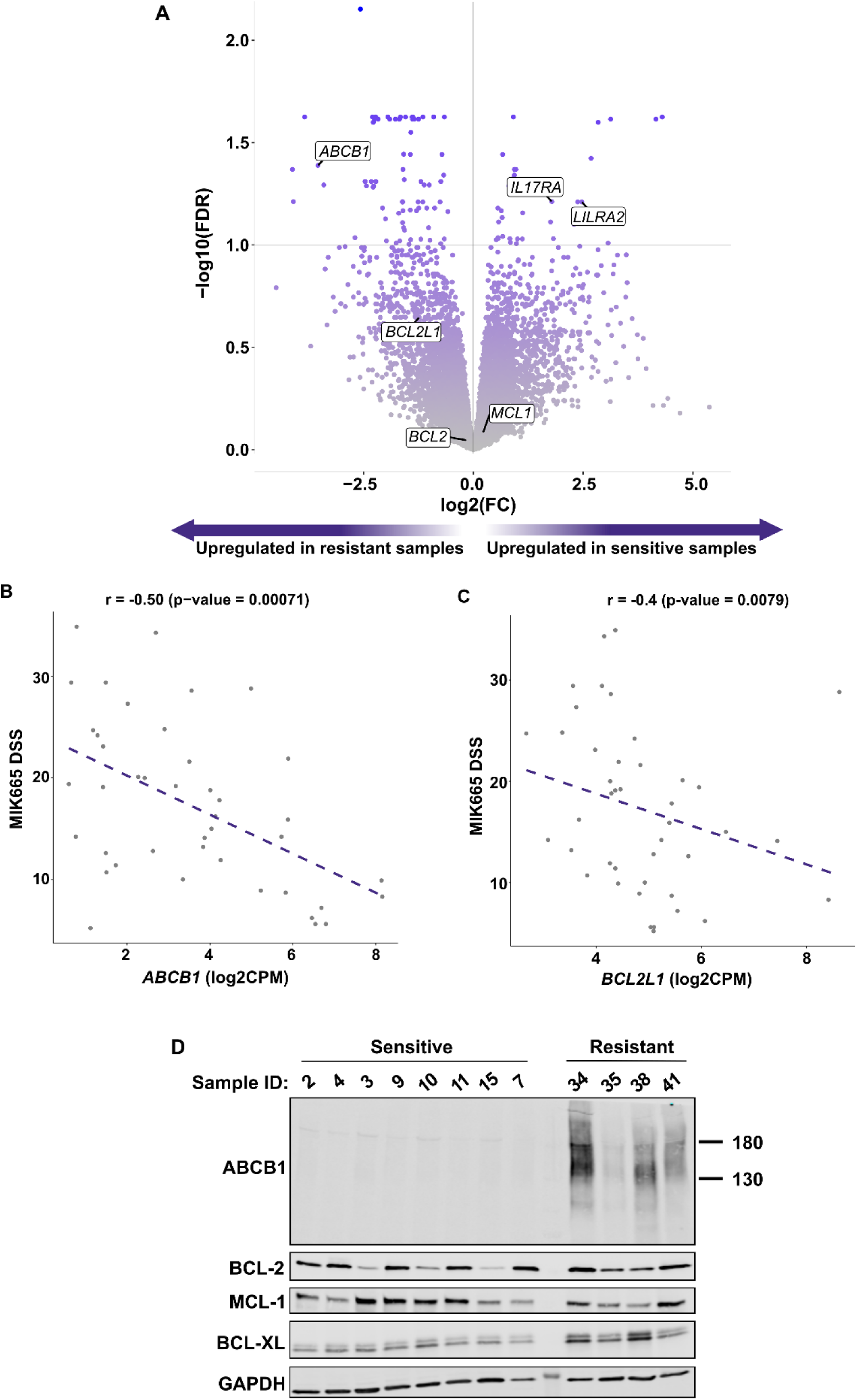
Differential gene expression analysis reveals markers of sensitivity and resistance to MIK665 in AML. **A)** Volcano plot showing genes analyzed in the differential gene expression analysis. Genes to the left of the vertical line represent those upregulated in the resistant group when compared to the sensitive group, whereas those to the right of the line are downregulated genes. Genes above the horizontal line (FDR cutoff at 0.1) are significantly differentially expressed (n=112). The labeled genes are *ABCB1* (enriched in the resistant samples; logFC = −3.54; FDR = 0.041), *LILRA2* (logFC = 2.47; FDR = 0.062) and *IL17RA* (logFC = 1.79; FDR = 0.061) (enriched in the sensitive samples), as well as *BCL2*, *MCL1*, and *BCL2L1*. The full list and statistics of the differentially expressed genes are available in Supplementary Table 7. **B)** Scatterplot showing an inverse correlation between *ABCB1* expression and MIK665 DSS in AML samples (n = 42; r = −0.5; p = 0.00071). **C)** Scatterplot showing an inverse correlation between *BCL2L1* and MIK665 DSS in AML samples (n = 42; r = −0.4; p = 0.0079). Correlations were performed using the Spearman method. **D)** Western blot showing the protein expression levels of ABCB1, BCL-2, MCL-1, and BCL-XL in a selection of MIK665-sensitive or resistant AML samples. log2FC: log2 fold change; log2CPM: log2 counts per million; FDR: false discovery rate.

*ABCB1* was among the most enriched genes in the resistant samples, with a log2 fold change of −3.54 (Figure 2A). Encoding the multi-drug resistance protein MDR1, also known as P-gp, *ABCB1* is associated with multi-class drug resistance in a range of cancers (28). *ABCB1* expression level was significantly upregulated in MIK665-resistant samples compared to both intermediate and sensitive samples (Supplementary Figure 6A). Across the samples, *ABCB1* expression was significantly inversely correlated to MIK665 DSS, indicating that *ABCB1* expression can be used as a marker of MIK665 resistance (Figure 2B). The difference in expression of *ABCB1* between the highest and lowest expressing samples was validated by RT-qPCR (Supplementary Figure 7, Supplementary Methods). To confirm ABCB1 expression at the protein level, we performed western blotting on a set of 12 samples (8 with low *ABCB1* and 4 with high *ABCB1* RNA expression). In concordance with the transcriptomic data, elevated levels of ABCB1 were detected in MIK665-resistant, but not in sensitive samples (Figure 2D, Supplementary Figure 8, Supplementary Methods).

Of the BCL-2 family members, *BCL2L1*, encoding the anti-apoptotic protein BCL-XL, had the highest expression in the resistant samples (Figure 2A, Supplementary Figure 6B). As with *ABCB1*, we detected a significant inverse correlation between *BCL2L1* expression and MIK665 DSS (Figure 2C). Interestingly, *BCL2L1* levels positively correlated with *ABCB1* levels in our sample cohort, a finding we confirmed in two publicly available datasets: BEAT and TCGA (Supplementary Figure 6C-E). Western blot analysis showed higher BCL-XL protein expression in MIK665-resistant compared to sensitive samples in our cohort. BCL-2 and MCL-1 levels varied across the sensitive and resistant samples, indicating that neither was linked to primary MIK665 resistance (Figure 2D, Supplementary Figure 8).

Together, these results indicate that high ABCB1 and high BCL-XL levels are useful indicators of MIK665 resistance and could constitute potential mechanisms of resistance to the drug in AML.

### Differentiation markers *LILRA2* and *IL17RA* are associated with MIK665 sensitivity

Differential gene expression analysis showed *LILRA2* and *IL17RA,* encoding leukocyte immunoglobulin-like and low affinity interleukin 17A receptors, respectively, to be among the top upregulated genes in MIK665-sensitive samples, with significantly higher expression levels compared to the intermediate and resistant samples (Figure 2A). In line with this finding, expression values of both genes were significantly correlated with MIK665 DSS values (Supplementary Figure 9A-D). Similar to *MCL1*, *LILRA2* and *IL17RA* are known to be associated with hematopoietic cell differentiation, notably monocytic and polymorphonuclear lineages (Supplementary Figure 10). To study the implications of this expression pattern on MIK665 sensitivity, we compared MIK665 response levels in our cohort across differentiation subtypes based on the French-American-British (FAB) classification. AML samples with a more differentiated cellular phenotype (FAB M4 or M5) were generally more sensitive to MIK665 compared to immature samples (FAB M0/M1 or M2), even though a small subset of less differentiated samples was also responsive (Supplementary Figure 9E). These findings suggest that more differentiated AML phenotypes, characterized by high *LILRA2* and *IL17RA* expression, are more likely to be sensitive to MCL-1 inhibition by MIK665, in line with our observation that MIK665 is more effective in monocytic populations.

### AML cell lines with high *ABCB1* expression demonstrate distinct dependency patterns on BCL-2 family members

To further investigate potential relationships between *ABCB1* and *BCL2* family anti-apoptotic genes, we analyzed gene expression (n=45) and dependency data (n=24) of AML cell lines (Figure 3A and 3B; Supplementary Figure 11). Cell lines were categorized as having a high (log2CPM > 2) or low (log2CPM < 2) *ABCB1* expression, and the two groups were contrasted for their expression of and dependence on *BCL2, MCL1,* and *BCL2L1*. AML cell lines with high *ABCB1* expression had significantly reduced *BCL2* and significantly increased *BCL2L1* expression compared to the cell lines with low *ABCB1* level, while no difference in the *MCL1* expression was detected between the two groups (Figure 3C). Concordantly, AML cell lines with high *ABCB1* expression demonstrated significantly higher dependency on *BCL2L1*, as well as significantly reduced *BCL2* and *MCL1* dependency (Figure 3D). Together, these findings suggest that AML cell lines exhibiting elevated *ABCB1* expression are more dependent on BCL-XL than on MCL-1 or BCL-2 for survival.

**Figure 3.**
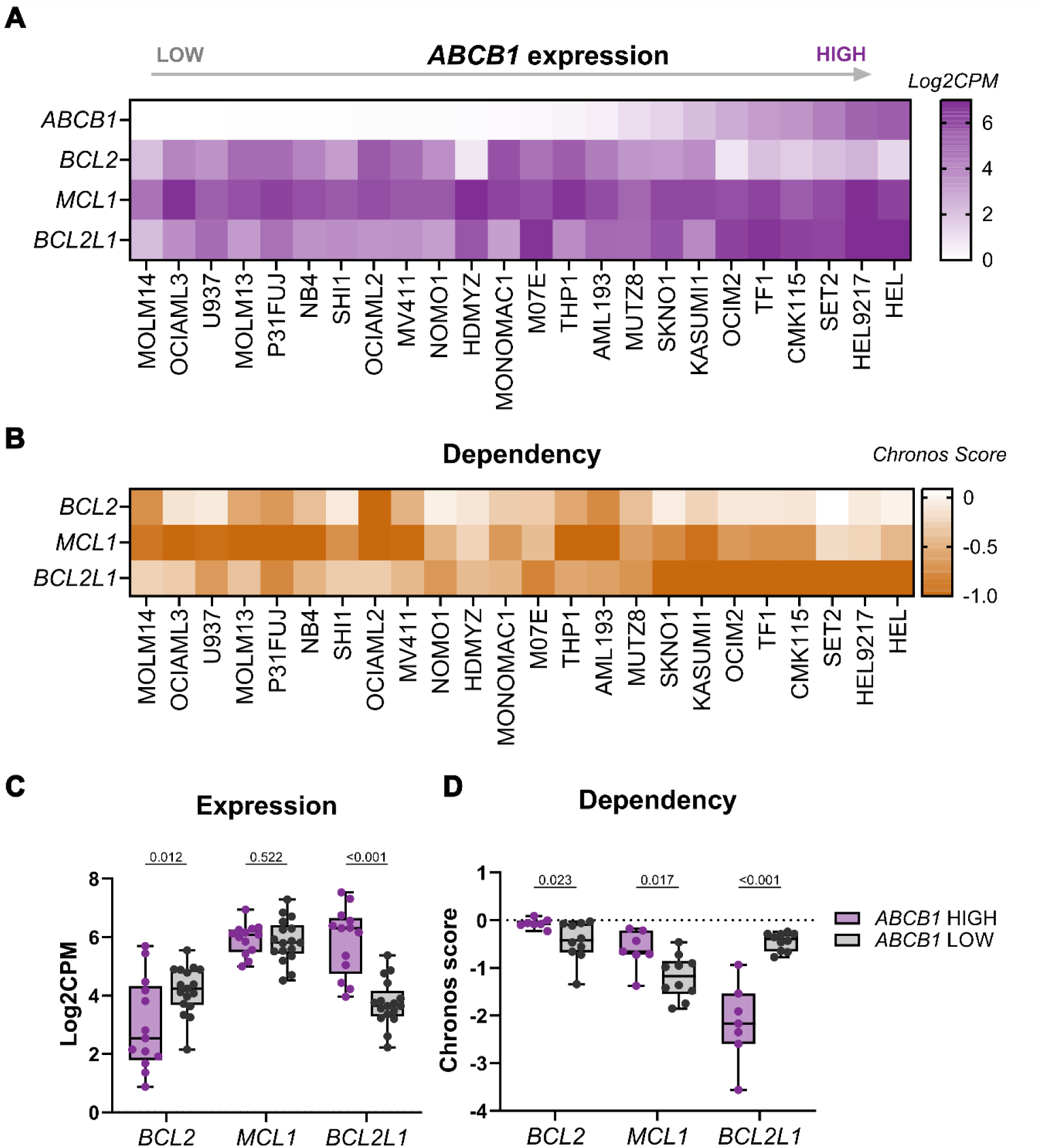
DepMap analysis of AML cell line data reveals that high *ABCB1* and low *MCL1* expression are associated with *BCL2L1* upregulation and dependency on it. A) Heatmap showing *ABCB1*, *BCL2*, *MCL1*, and *BCL2L1* expression (log2CPM) in AML cell lines sorted based on *ABCB1* expression (log2CPM). **B)** Heatmap showing dependency (Chronos score) of AML cell lines on *BCL2*, *MCL1*, and *BCL2L1.* Cell line order is based on *ABCB1* expression. Boxplots representing *BCL2*, *MCL1,* and *BCL2L1* C) expression and **D)** dependency in cell lines with high (purple) and low (grey) *ABCB1* levels. Log2CPM ≥ 2 was defined as a high expression of *ABCB1*, whereas log2CPM < 2 was considered low expression. Each dot represents a single cell line. Significance was evaluated using the 2-sample t-test. log2CPM: log2 counts per million.

### *ABCB1* expression is a strong predictive biomarker of MIK665 resistance

To evaluate the predictive effect of *ABCB1* expression on MIK665 response in AML samples, we plotted the distribution of *ABCB1* expression values across the response subgroups in our cohort. The density curves of expression per subgroup showed distinct peaks, with the biggest separation difference between the resistant subgroup compared to the sensitive and intermediate subgroups (Figure 4A, Supplementary Figure 6A). We then performed a receiver operating characteristics (ROC) curve analysis to assess the ability of *ABCB1* expression to distinguish MIK665-resistant from sensitive and intermediate samples. The cutoff with the optimal classifying performance for *ABCB1* expression was 5.22 (log2CPM) as identified using the Youden index. This resulted in an area under the ROC curve of 0.856, with a sensitivity of 80%, a specificity of 90.6%, and a positive predictive value of 72.7% (Figure 4B-C).

**Figure 4.**
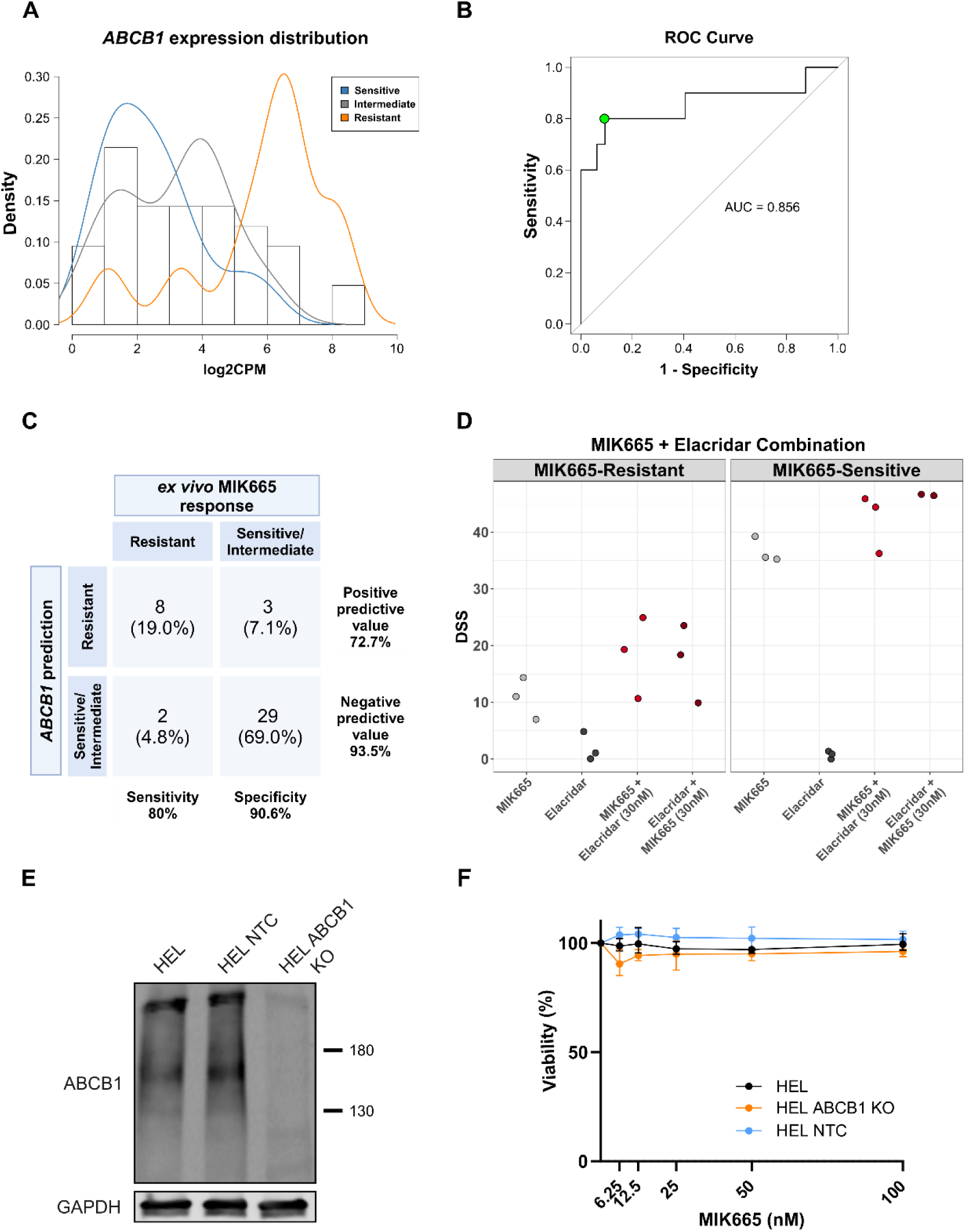
ABCB1 inhibition in AML with high *ABCB1* expression is predictive of MIK665 resistance. **A)** Density curves representing the distribution of *ABCB1* expression values across the MIK665 response groups. **B)** Receiver operator characteristic ROC curve of *ABCB1* expression as a predictor of MIK665 resistance. The optimal cutoff value for *ABCB1* was selected using the Youden index and is indicated by the green dot. **C)** Contingency matrix showing the performance statistics of the test when using an *ABCB1* cutoff value of 5.22. Created with BioRender.com. **D)** Dotplot showing the DSS of MIK665 and ABCB1 inhibitor elacridar alone or in combination in AML samples with MIK665 resistance (high *ABCB1*: log2CPM > 6) or MIK665 sensitivity (low *ABCB1*: log2CPM < 6). Grey dots represent the DSS values of the single agents, whereas red dots represent the DSS values of the combination (where one drug is increased along its concentration range while the other is fixed at 30 nM). Significance was evaluated using the paired sample t-test. **E)** Western blot showing the protein expression level of ABCB1 in HEL ABCB1 knockout (HEL ABCB1 KO) cells, as compared to parental and non-target control (NTC) cells. **F)** Dose-response curves of HEL, HEL ABCB1 knockout and HEL non-target control cells when treated with MIK665 (N=3). Error bars represent the standard deviation. All responses were measured using CellTiter Glo following a 48 h incubation with the drugs. log2CPM: log2 counts per million; AUC: area under the curve.

### ABCB1 inhibition does not overcome MIK665 resistance

As ABCB1 is a known driver of multidrug resistance across several cancer types, we sought to evaluate its role as a potential target in MIK665 resistant AML. We selected three ABCB1 high (MIK665 DSS < 15) and three ABCB1 low (MIK DSS > 35) samples and tested these with the ABCB1 inhibitor elacridar in combination with MIK665. The combination of elacridar and MIK665 did not result in significantly higher responses in the ABCB1 high samples, compared to either agent alone (Figure 4D). To further explore if ABCB1 inhibition can restore sensitivity to MIK665, we tested the MIK665 and elacridar combination in the HEL AML cell line. The HEL cell line was selected due to its high *ABCB1* expression level and was therefore expected to be resistant to MCL-1 inhibition by MIK665 (Figure 3A). However, HEL cells did not respond to either drug individually or in combination, indicating that chemical inhibition of ABCB1 cannot restore sensitivity to MCL-1 inhibition by MIK665. To mechanistically investigate the role of ABCB1 in MIK665 resistance, we generated a HEL ABCB1 knockout cell line using CRISPR-Cas9 (Figure 4E). Upon testing of MIK665 in the HEL ABCB1 knockout cells, drug sensitivity was not increased when compared with HEL parental and HEL non-target control (Figure 4F). Overall, we conclude that ABCB1 targeting cannot reverse MIK665 resistance, however, elevated *ABCB1* expression serves as a predictive biomarker of MIK665 resistance in AML.

### MIK665 combined with venetoclax is an effective combination in AML samples with primary resistance to either of the single agents

To evaluate whether co-targeting other anti-apoptotic proteins is effective in MIK665-resistant samples, we tested combinations of MIK665 with BCL-XL inhibitor A-1331852 or BCL-2 inhibitor venetoclax. The combination of MIK665 and A-1331852 did not result in increased sensitivity of the MIK665-resistant samples compared to treatment with MIK665 alone, indicating that co-targeting of BCL-XL and MCL-1 does not restore MIK665 sensitivity (Supplementary Figure 12). However, all three MIK665-resistant samples had significantly higher DSS values to the MIK665 and venetoclax combination compared to MIK665 alone, which suggest that targeting both MCL-1 and BCL-2 is an effective strategy to overcome resistance to MIK665 (Figure 5A). Interestingly, we noticed that one of the MIK665-resistant samples that responded well to the combination also had *ex vivo* primary resistance to venetoclax. To further assess the potential use of this combination in the context of primary venetoclax resistance, we tested three additional samples with primary venetoclax resistance (Supplementary Figure 5, Supplementary Table 6). The combination showed significantly higher DSS values in all three samples compared to venetoclax alone (Figure 5A). The venetoclax dose-response curves with MIK665 fixed at 30 nM further illustrated the benefit of the addition of MIK665 to venetoclax in these samples (Figure 5B). Together, these results demonstrate that a combination of MIK665 and venetoclax is an efficacious strategy to overcome primary resistance to either of the two agents.

**Figure 5.**
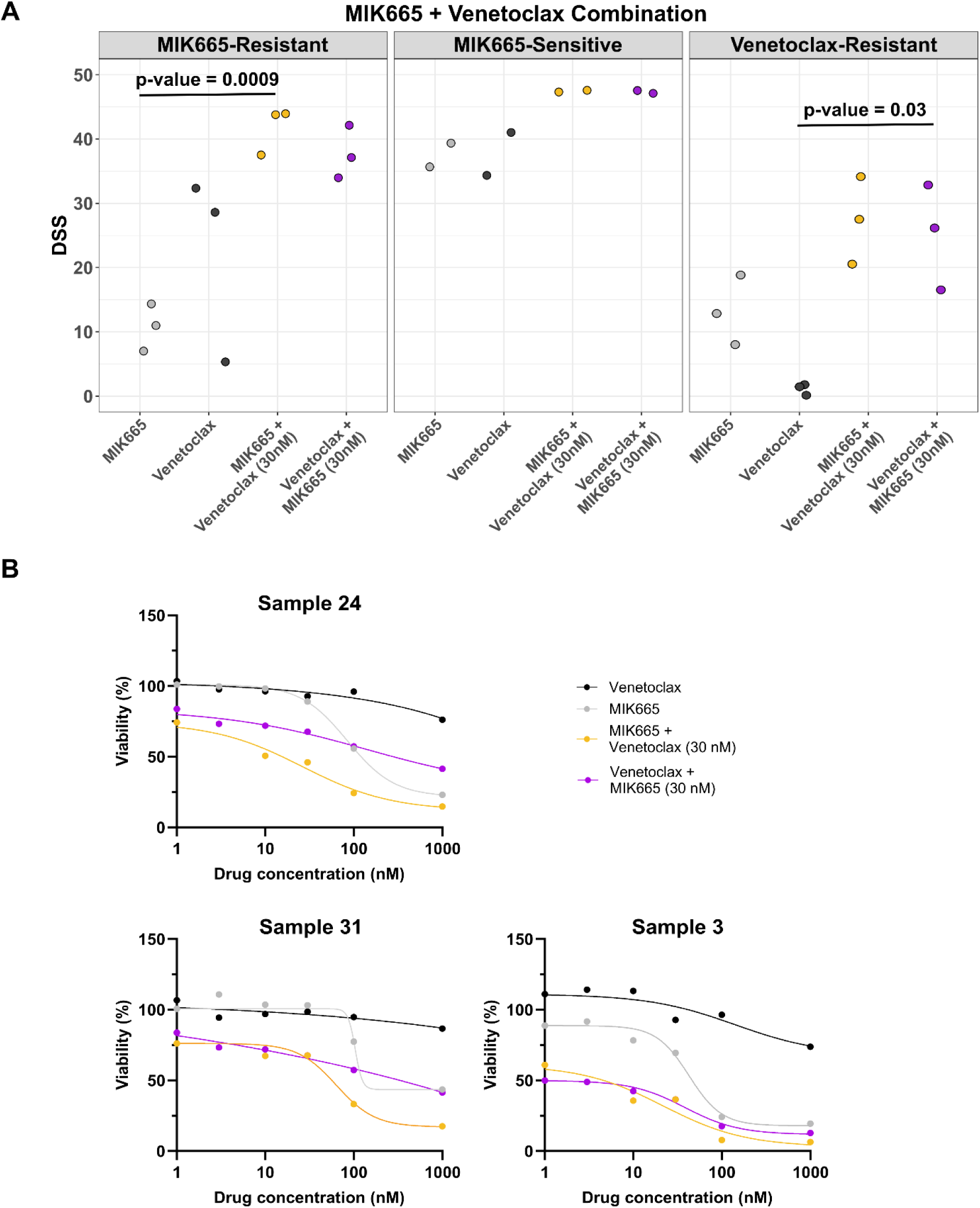
A combination of MIK665 and BCL-2 inhibitor venetoclax is more effective compared to each agent alone in selected AML samples. **A)** Results showing the DSS of MIK665 and venetoclax alone or in combination in MIK665-resistant, MIK665-sensitive and venetoclax-resistant AML samples. MIK665-resistant samples were selected as having high *ABCB1* expression (log2CPM > 6) and a MIK665 DSS < 15. MIK665-sensitive samples were selected as having low *ABCB1* expression (log2CPM < 6) and a MIK665 DSS > 30. Venetoclax-resistant samples were selected as having a venetoclax DSS < 10. Grey dots represent the DSS values of the single agents, whereas orange or purple dots represent the DSS values of the combinations (where one drug is increased along its concentration range while the other is fixed at 30 nM). All responses are measured following a 48 h incubation with the drugs. Significance was evaluated using the paired sample t-test. **B)** Dose response curves of the 3 venetoclax-resistant patient samples as tested in panel A.

### MIK665 combined with venetoclax restores sensitivity in AML cell lines with acquired venetoclax resistance

As acquired resistance to venetoclax is commonly observed following venetoclax-based treatment, we tested the MIK665 and venetoclax combination in AML cell line models with acquired venetoclax resistance. We generated VenR cell lines MV4-11_VenR, Kasumi-1_VenR, MOLM-13_VenR and HL-60_VenR by exposing parental cell lines to increasing concentrations of venetoclax. In all VenR cell lines, MIK665 in combination with venetoclax restored sensitivity. The combination showed high synergy in all cell lines, with a greater effect in MV4-11_VenR and MOLM-13_VenR (Figure 6A). Comparison of BCL-2 family protein levels of parental and VenR cell lines showed a trend towards a decrease in BCL-2 expression and an increase in BCL-XL expression upon acquired venetoclax resistance (Figure 6B). These findings show that MIK665 restores sensitivity to venetoclax in AML with acquired venetoclax resistance, indicating the potential clinical relevance of this combination upon patient relapse to venetoclax-based treatment regimens.

**Figure 6.**
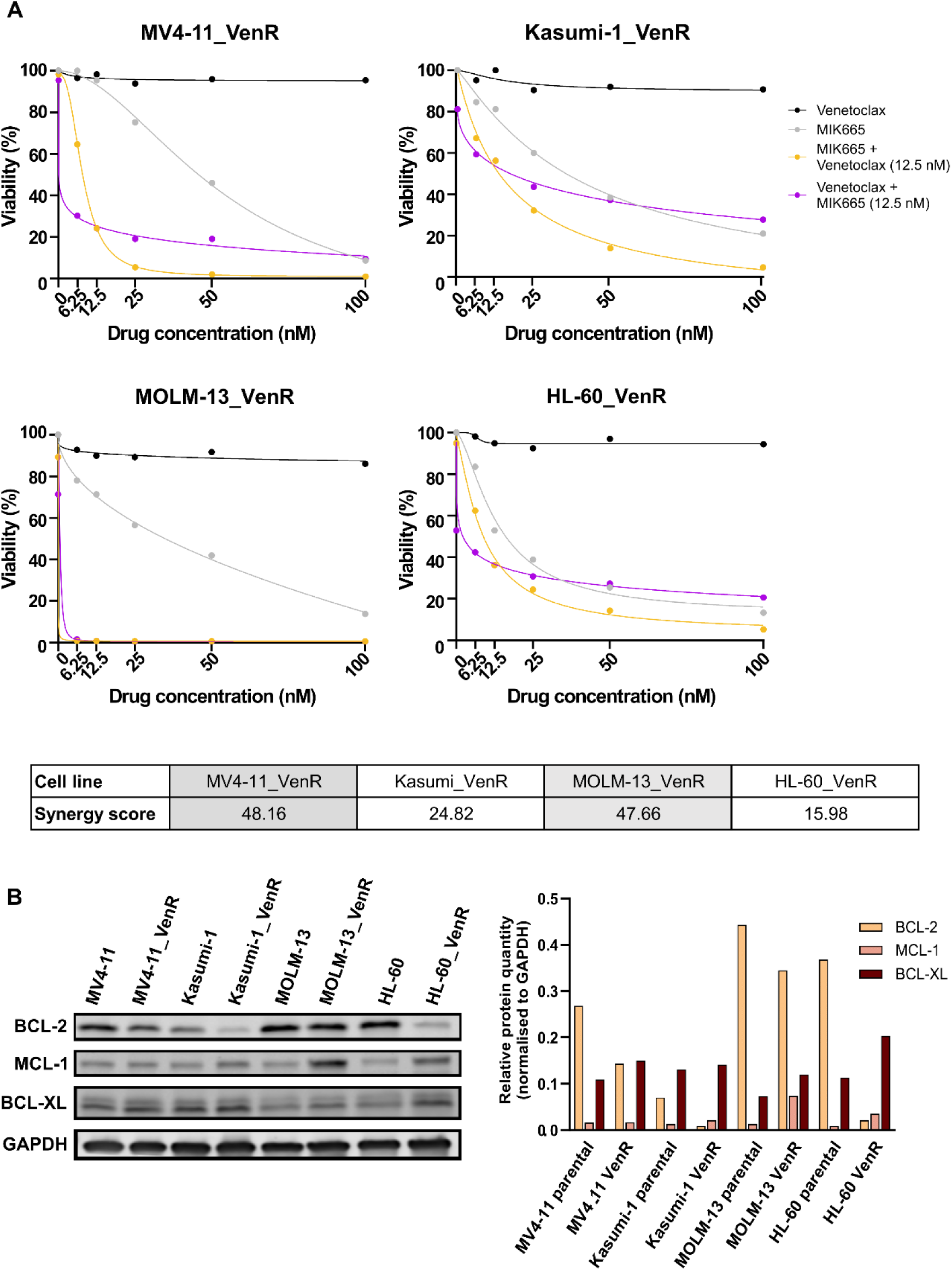
A combination of MIK665 and venetoclax is effective in AML cell lines with acquired resistance to venetoclax. **A)** Dose-response curves of MIK665 and venetoclax alone and in combination in MV4-11 venetoclax-resistant (MV4-11_VenR), Kasumi-1 venetoclax-resistant (Kasumi-1_VenR), MOLM-13 venetoclax-resistant (MOLM-13_VenR) and HL-60 venetoclax-resistant (HL-60_VenR) cell lines, where one drug is increased along its concentration range while the other is fixed at 12.5 nM. Responses are measured by CellTiter-Glo following a 48 h incubation. ZIP synergy scores for the combination in each of the cell line are reported in the table. **B)** Western blot showing the protein expression level of BCL-2, MCL-1, and BCL-XL in parental and VenR cell lines, and the protein quantification normalized to the respective GAPDH levels.

## Discussion

Our assessment of MCL-1 inhibitor MIK665 activity in primary AML patient samples resulted in varied responses, reflecting the heterogeneous response observed in clinical trials (14,15). *ABCB1* was among the most significantly upregulated genes in the MIK665-resistant samples, and logistic regression and ROC curve analyses revealed a strong performance of *ABCB1* expression in identifying MIK665-resistant samples. Nonetheless, direct ABCB1 inhibition did not reverse MIK665 resistance. Instead, co-inhibition of MCL1 and BCL2 by the addition of venetoclax proved to be an effective strategy in the MIK665-resistant population.

ABCB1, commonly referred to as multidrug resistance protein 1 (MDR1) or P-glycoprotein (P-gp), can confer resistance to several cancer therapeutics (29). Our previous research has demonstrated that ABCB1-mediated drug efflux represents a resistance mechanism to MCL-1 inhibitors in multiple myeloma (30). In AML, the chemotherapeutic daunorubicin is a known ABCB1 substrate, and high *ABCB1* expression is enriched in adverse European LeukemiaNet (ELN) risk subgroups with poor outcome in patients treated with standard chemotherapy (31–33). To determine whether ABCB1 inhibition can overcome MIK665 resistance in AML, we evaluated the efficacy of a combination of MIK665 and third generation ABCB1-specific inhibitor elacridar in samples with high ABCB1 expression and low response to MIK665. However, the combination was not more effective compared to MIK665 alone. This is in line with previous studies showing that ABCB1 expression associates with resistance to intensive chemotherapy in AML patients, but that its co-inhibition does not improve prognosis (33). Furthermore, knockout of ABCB1 using CRISPR-Cas9 in AML cells having a high basal level of *ABCB1* expression and resistance to MIK665, did not lead to an increased sensitivity. Together, these data indicate that while ABCB1 does not represent a suitable target to overcome MIK665 resistance, high *ABCB1* expression can be used as a biomarker of *ex vivo* MIK665 resistance in AML.

We found *BCL2L1,* a gene encoding the anti-apoptotic protein BCL-XL, to be significantly correlated with ABCB1 expression in three AML patient cohorts. Previous research has reported that high levels of BCL-XL correlate with resistance to MCL-1 inhibition (13,27,34), a finding also observed in this study’s MIK665-resistant patient samples. AML cell line analysis further demonstrated that high *ABCB1* expression had significantly higher expression of and dependence on *BCL2L1.* Kuusanmäki et al. found that high BCL-XL expression was correlated with sensitivity to BCL-XL inhibitors in erythroleukemia (FAB M6/M7) (34). However, in our study MIK665-resistant samples did not respond to the BCL-XL inhibitor A1331852 alone or in combination with MIK665. This is likely due to our cohort lacking samples with M6/M7 phenotypes. Further analysis indicated that *ABCB1* and *BCL2L1* were significantly enriched in FAB types M0/M1 and M2 compared to M4 and M5 (Supplementary Figure 13, Supplementary Table 8). Supporting this, samples in our cohort with higher expression levels of *LILRA2* and *ILI7RA*, markers of phenotypically mature monocytic-like AML subtypes (notably M4 and M5), were more sensitive to MCL-1 inhibition by MIK665 (35). These results align with earlier studies demonstrating that AML FAB subtypes M4 and M5 have higher expression of and dependence on MCL-1 and are thus sensitive to MCL-1 inhibition (36,37).

*In vitro* studies have indicated that MCL-1 inhibitors synergize with BCL-2 inhibitors, namely venetoclax, leading to the evaluation of this combination for AML patients in several clinical trials. (11,13,15). Aligning with these observations, we found that the combination of MIK665 and venetoclax is effective in samples with MIK665 primary resistance. Interestingly, this combination was also effective in AML samples with primary venetoclax resistance, a finding with important clinical relevance, since up to 30% of AML patients treated with venetoclax-based regimens are found to be primary refractory to the treatment (2,38–41). Previous clinical trials have reported cases of disease progression and acquired resistance to venetoclax-based therapies following initial remissions in over 50% of patients until time last of follow-up, leaving them with limited treatment options and a dismal prognosis (40,42). Using AML cell lines with acquired venetoclax resistance as a model for clinical venetoclax relapse, we observed that the MIK665 and venetoclax combination was also highly effective in this context. This is in line with previous findings showing that MCL1-upregulation contributes to acquired venetoclax resistance, and that the addition of an MCL-1 inhibitor has the potential to restore sensitivity (43,44). Overall, the combination of MIK665 and venetoclax was effective in AML patient samples with primary resistance to either of the single agents, as well as AML cell lines with acquired venetoclax resistance, a finding warranting further clinical evaluation. The elevated sensitivity of such AML patient subgroups to the combination of MIK665 and venetoclax suggests that lower treatment doses could be administered, thereby improving toxicity profiles, which have so far represented a major obstacle for MCL-1 inhibitor development.

In summary, the associations between diverse molecular features of AML samples and their sensitivity to MIK665 described in this study provide valuable insights from a precision medicine and clinical trial design viewpoint. Our study identifies a sizable target AML patient population with differentiated disease that is susceptible to MCL-1 inhibition by MIK665. Moreover, we show that elevated *ABCB1* expression represents a predictive biomarker of resistance to MIK665, which can be overcome by the addition of venetoclax. Our findings support the evaluation of the combination of MIK665 with venetoclax in resistant patient populations to restore and prolong clinical responses.

## Data Availability

All data produced in the present work are contained in the manuscript and its supplementary data and are available upon reasonable request to the corresponding author.

## Acknowledgements

The authors of this work would like to thank the Finnish Hematology Registry and Clinical Biobank for providing the patient samples and data, the contributing clinicians and nurses, and especially the donors. The authors also acknowledge the FIMM High Throughput Biomedicine Unit for the drug plate preparation, as well as the FIMM Technology Center sequencing and bioinformatics units for preparing and curating the samples’ molecular profiles. The units are hosted by the University of Helsinki and are supported by HiLIFE and Biocenter Finland. This work is expected to contribute to the fulfillment of the PhD requirements of J.S. and R.N., both doctoral candidates at the University of Helsinki. The authors would like to extend further thanks to Minna Suvela and Siv Knaappila for processing the samples used in the study, and to Lisa Eick for providing input into the biomarker analysis. The authors also appreciate the valuable input and revisions received from colleagues at FIMM, Novartis, the Helsinki University Hospital and Comprehensive Cancer Center and Servier Laboratories. This work was supported by funding received by C.A.H. from Novartis, the Research Council of Finland (grant no. 334781, 352265, 357686, and 320185), the Sigrid Jusélius Foundation, and the Cancer Foundation Finland.

## Author contributions

J.S., R.N., E.K., S.A., H.K., M.T., A.P., J.M. and K.K. designed and performed the experimental work. J.S., R.N., E.K., N.I., and E.D. carried out the result analysis and interpretation. J.S. and R.N. drafted the manuscript, and all authors contributed to its revision. C.A.H. and E.H. conceived and supervised the study. C.A.H. provided infrastructure to carry out the work.

