## Supplementary Figures and Methods for "Predictors of Response and Rational Combinations for the Novel MCL-1 Inhibitor MIK665 in Acute Myeloid Leukemia"

| | 1 | 2 | 3 | 4 | 5 | 6 | 7 | 8 | 9 | 10 | 11 | 12 |
|---|---|---|---|---|---|---|---|---|---|---|---|---|
| A | empty | MIK665_1000 | Venetoclax_1 | DMSO | A1331852_30<br>MIK665_3 | MIK665_30<br>A133852_3 | Venetoclax_30<br>MIK665_30 | MIK665_30<br>Venetoclax_10 | BzCl | MIK665_30<br>Elacridar_100 | DMSO | empty |
| B | MIK665_1 | A1331852_1 | Venetoclax_3 | Elacridar_3 | A1331852_30<br>MIK665_10 | MIK665_30<br>A133852_10 | BzCl | MIK665_30<br>Venetoclax_100 | Elacridar_30<br>MIK665_100 | MIK665_30<br>Elacridar_1000 | | |
| C | MIK665_3 | BzCl | Venetoclax_10 | Elacridar_10 | DMSO | MIK665_30<br>A133852_100 | Venetoclax_30<br>MIK665_100 | MIK665_30<br>Venetoclax_1000 | DMSO | | | DMSO |
| D | MIK665_10 | A1331852_3 | Venetoclax_30 | Elacridar_30 | A1331852_30<br>MIK665_30 | MIK665_30<br>A133852_1000 | DMSO | Elacridar_30<br>MIK665_1 | Elacridar_30<br>MIK665_1000 | | | |
| E | DMSO | A1331852_10 | Venetoclax_100 | BzCl | A1331852_30<br>MIK665_100 | Venetoclax_30<br>MIK665_1 | Venetoclax_30<br>MIK665_1000 | Elacridar_30<br>MIK665_3 | MIK665_30<br>Elacridar_1 | DMSO | | BzCl |
| F | MIK665_30 | A1331852_30 | Venetoclax_1000 | Elacridar_100 | A1331852_30<br>MIK665_1000 | DMSO | MIK665_30<br>Venetoclax_1 | BzCl | MIK665_30<br>Elacridar_3 | | | |
| G | MIK665_100 | A1331852_100 | DMSO | Elacridar_1000 | MIK665_30<br>A133852_1 | Venetoclax_30<br>MIK665_3 | MIK665_30<br>Venetoclax_3 | Elacridar_30<br>MIK665_10 | MIK665_30<br>Elacridar_10 | | BzCl | |
| H | empty | A1331852_1000 | Elacridar_1 | A1331852_30<br>MIK665_1 | BzCl | Venetoclax_30<br>MIK665_10 | DMSO | Elacridar_30<br>MIK665_30 | DMSO | | | empty |

**Supplementary Figure 1. Combination drug plate layout.** Layout of drugs and drug concentrations in a 96-well plate used for the patient sample drug combination tests. The inhibitors were each tested either as single agents in 6 increasing concentrations (range of 1 to 1000 nM) or combined with another drug fixed at 30 nM. The concentrations are indicated after the drug name.

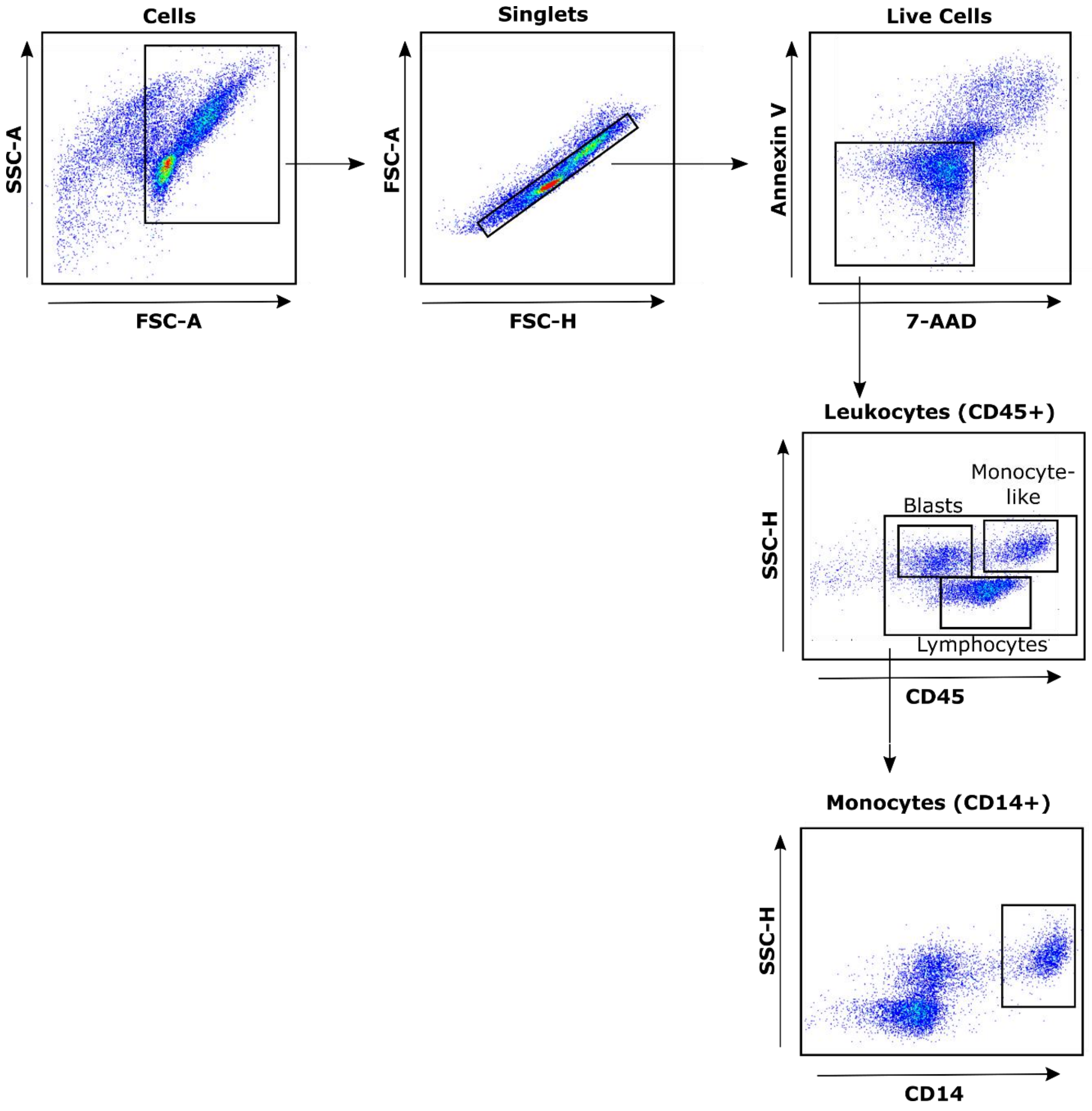


**Supplementary Figure 2. Gating strategy.** The flow cytometry gating strategy used to distinguish different constituent cell types in the AML patient sample cohort.

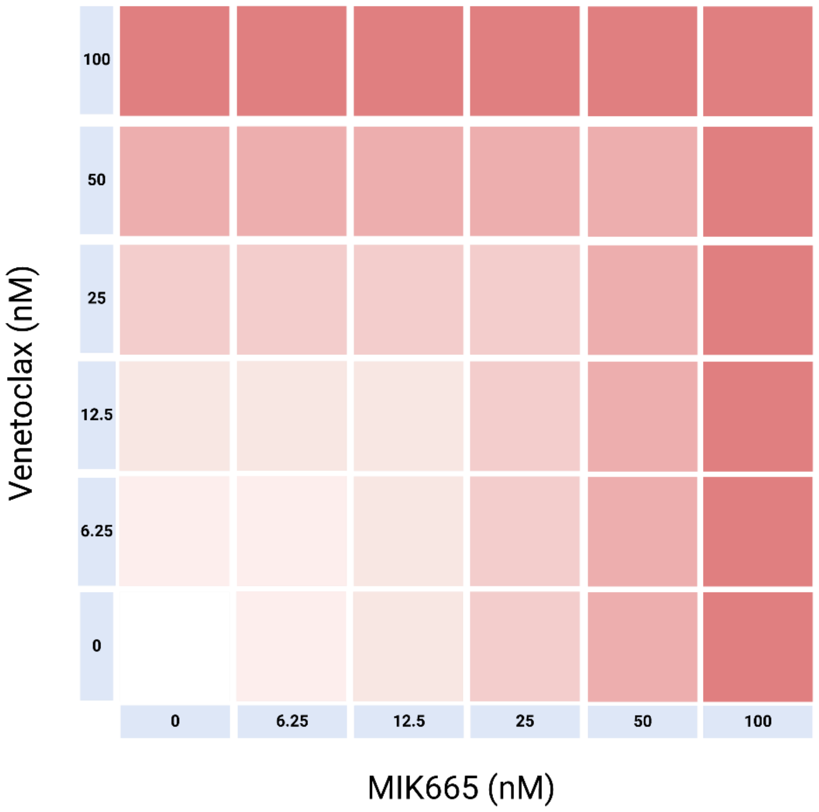


**Supplementary Figure 3. Schematic of the drug combination tested with MV4-11_VenR, Kasumi-1_VenR, MOLM-13_VenR and HL-60_VenR cell lines.** Each drug was tested across an increasing 5-concentration range from 6.25 to 100 nM either alone or in combination. Created with BioRender.com.

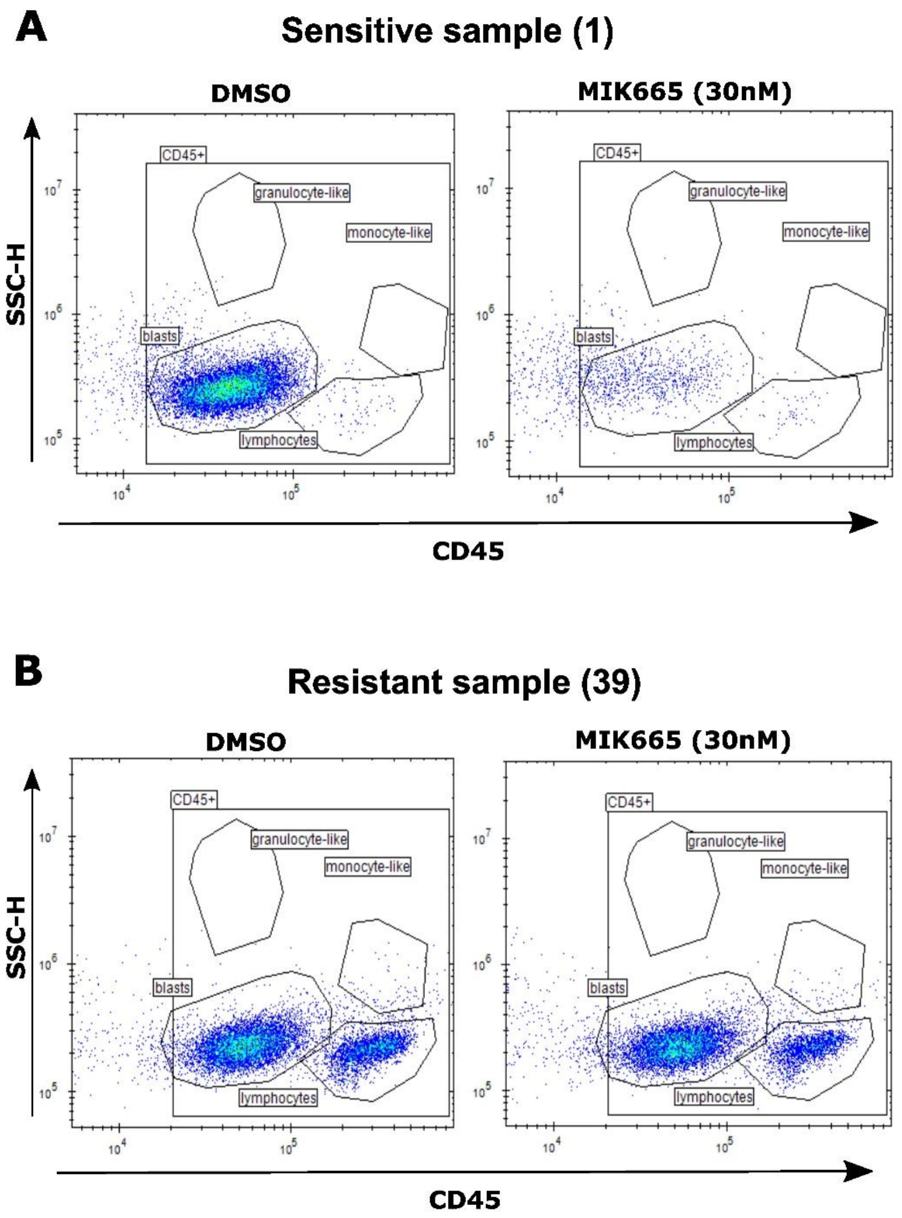


**Supplementary Figure 4. Representative dot plots showing the response of a sensitive and resistant AML sample to MIK665. A)** A sensitive sample (sample ID:1) shows a decrease in blast cells upon MIK665 treatment when compared with the DMSO control. **B)** A resistant sample (sample ID: 39) shows no change in blast cells upon MIK665 treatment when compared to DMSO, indicating the resistance of the sample to the treatment. Following 48 h incubation with MIK665 (30 nM) samples were analyzed using multi-parametric flow cytometry.

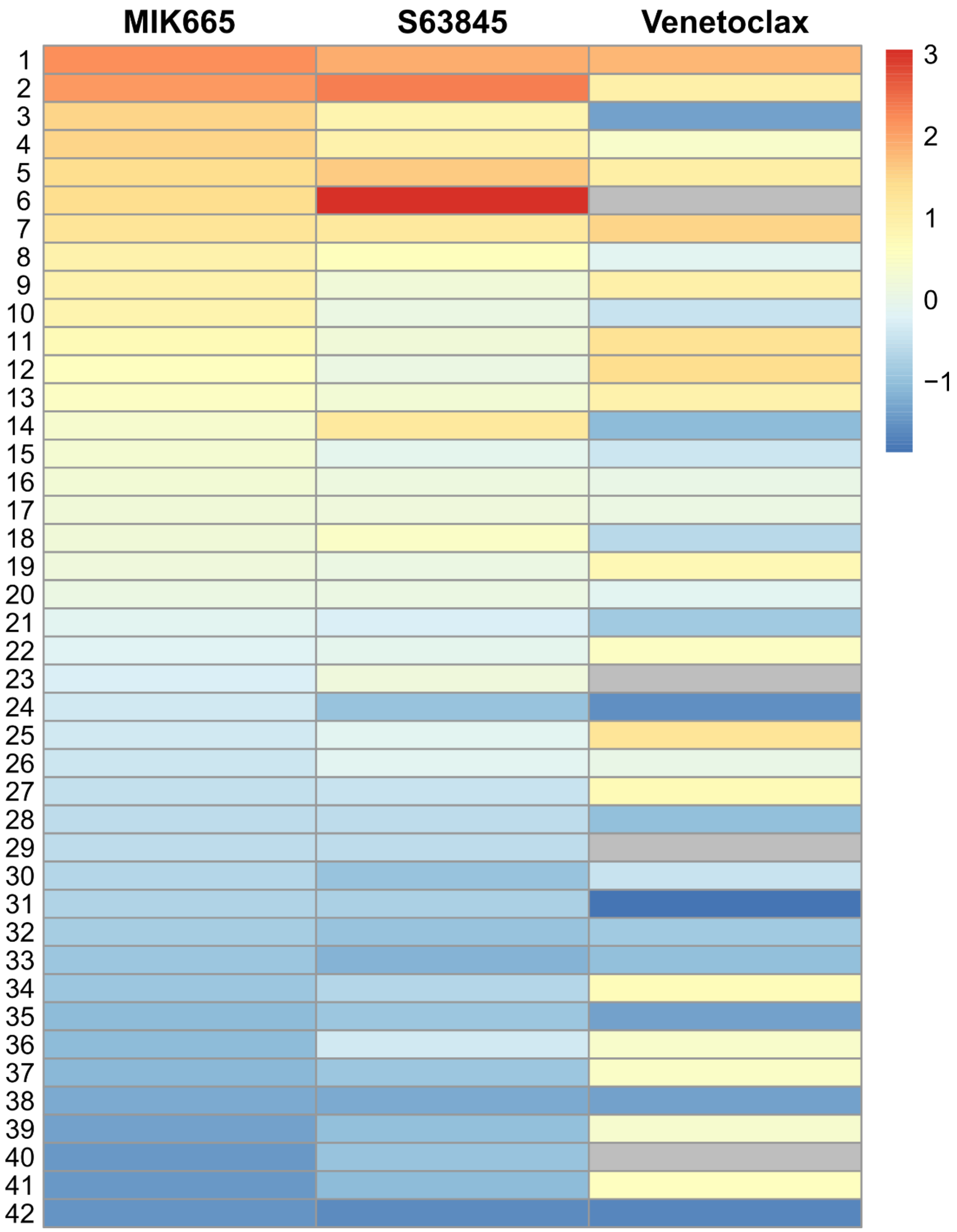


**Supplementary Figure 5. Heatmap summarizing the response of the AML sample cohort to MCL-1 inhibitors MIK665 and S63845, and to BCL-2 inhibitor venetoclax.** The DSS values obtained for the leukocyte populations were scaled and centered by drug, with low DSS values (blue) corresponding to drug resistance, and high DSS values (red) corresponding to drug sensitivity. Samples are ordered on the y-axis by MIK665 sensitivity. Grey boxes indicate missing or censored values.

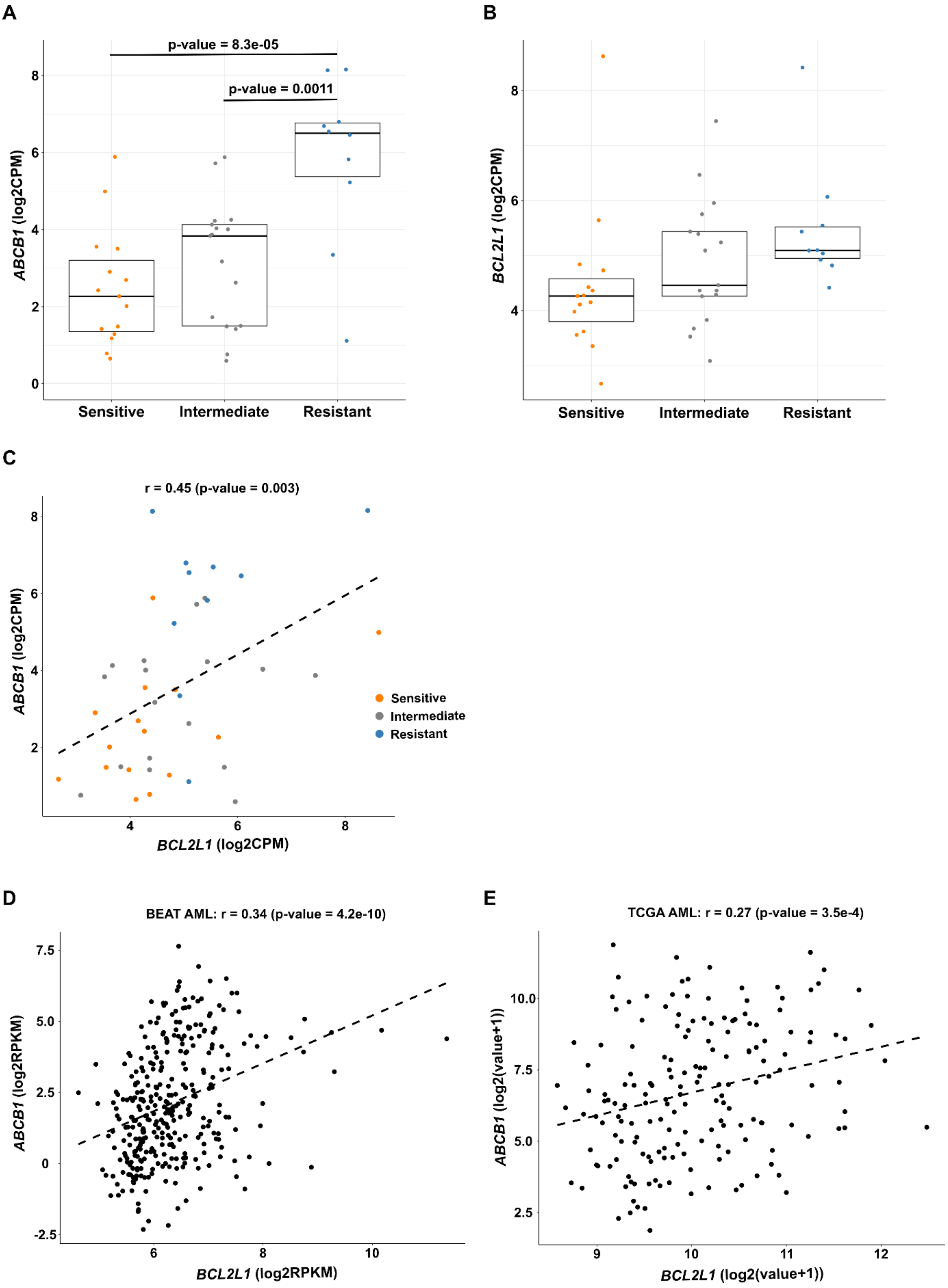


**Supplementary Figure 6. *ABCB1* and *BCL2L1* have higher gene expression levels in the MIK665-resistant sample subgroup. A)** The median level of expression of *ABCB1* increases significantly with MIK665 resistance (anova p-value = 8.4E-5), and it is significantly higher in resistant samples compared to sensitive and intermediate MIK665 response groups, by the Tukey test. **B)** The median gene expression level of *BCL2L1* shows an increasing trend from MIK665-sensitive to resistant samples. *ABCB1* and *BCL2L1* expressions correlate positively and significantly in primary patient samples from the **C)** FIMM, **D)** BEAT, and **E)** TCGA AML cohorts, using the Person correlation method.

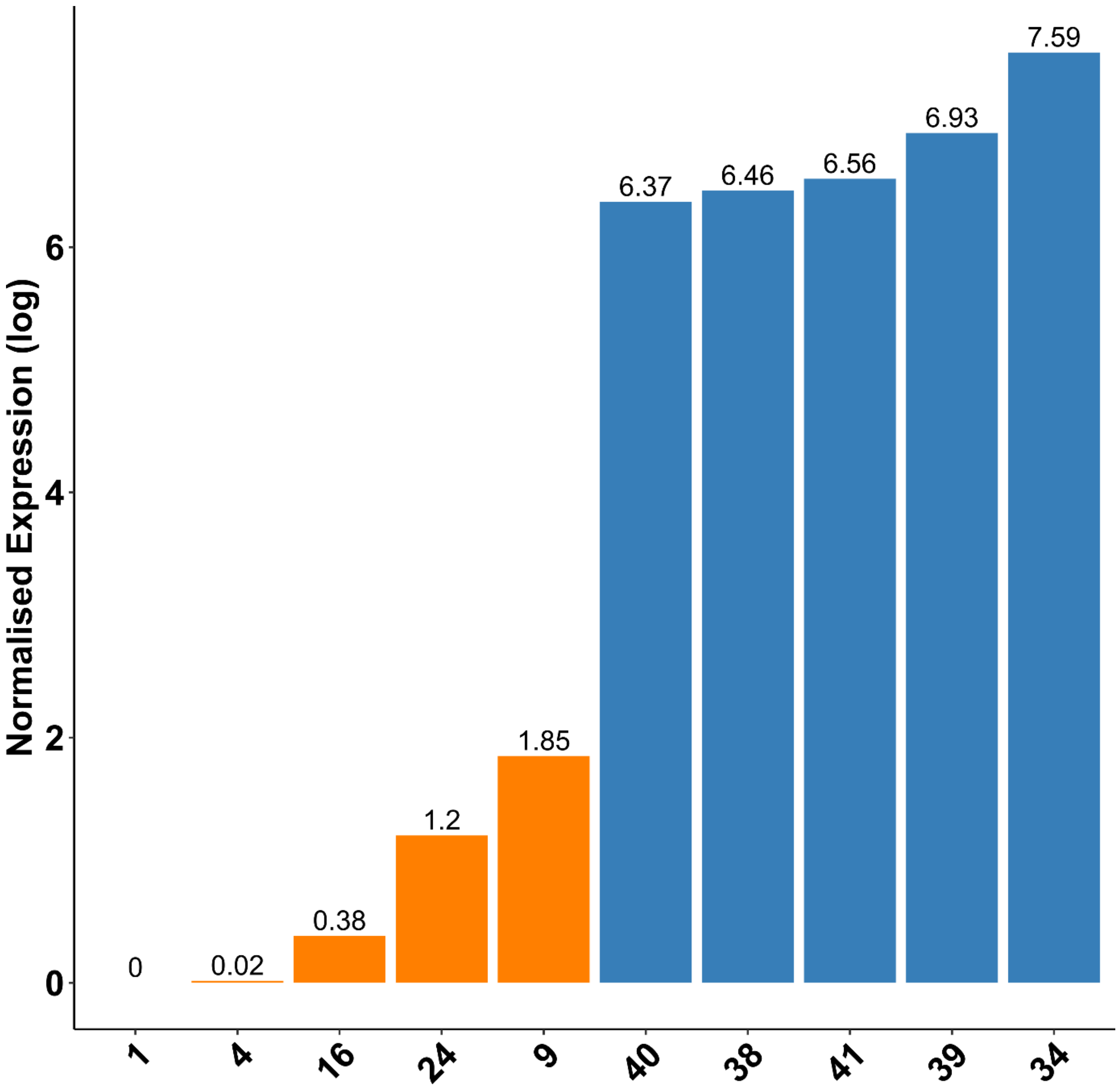


**Supplementary Figure 7. RT-qPCR experiments verify RNA sequencing data.** To validate *ABCB1* expression levels obtained from RNA sequencing data, confirmatory RT-qPCR was performed on 10 primary patient samples: 5 with low (orange) and 5 with high (blue) *ABCB1* expression. The computed normalized log fold-change ratios per sample are shown above each bar. The housekeeping genes used were *NONO*, *HNRNPC*, *EIF4B*.

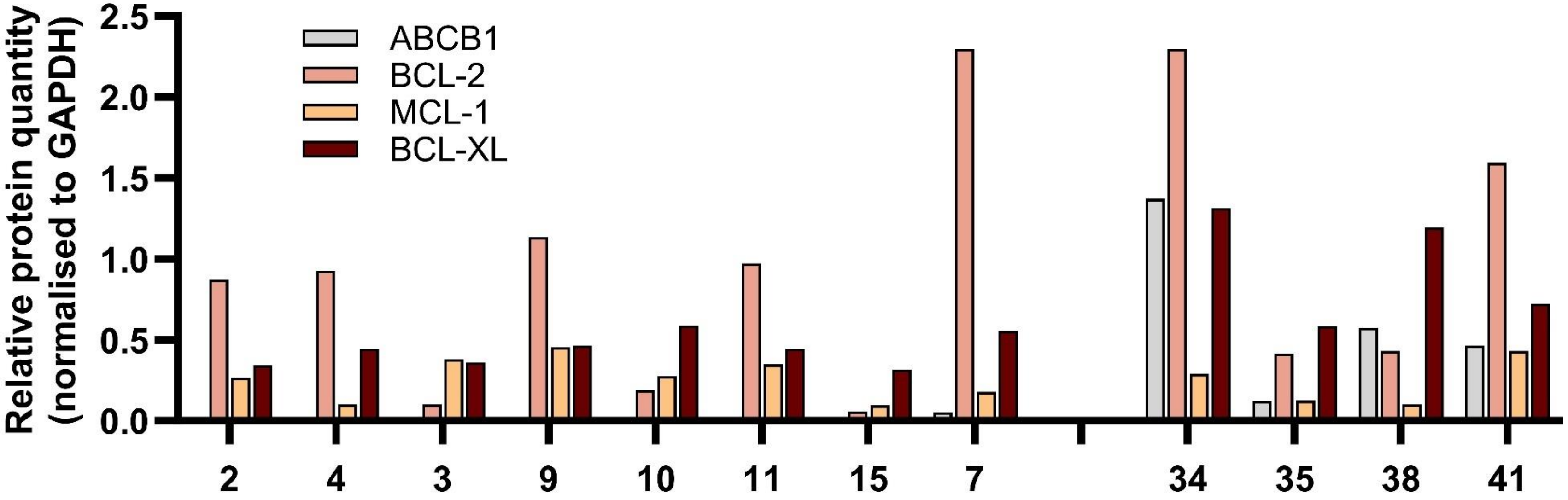


**Supplementary Figure 8. Quantification of western blots measuring ABCB1, BCL-2, MCL-1, and BCL-XL levels in MIK665-sensitive and resistant patient samples.** The relative protein quantity for each sample is normalized relative to GAPDH protein expression.

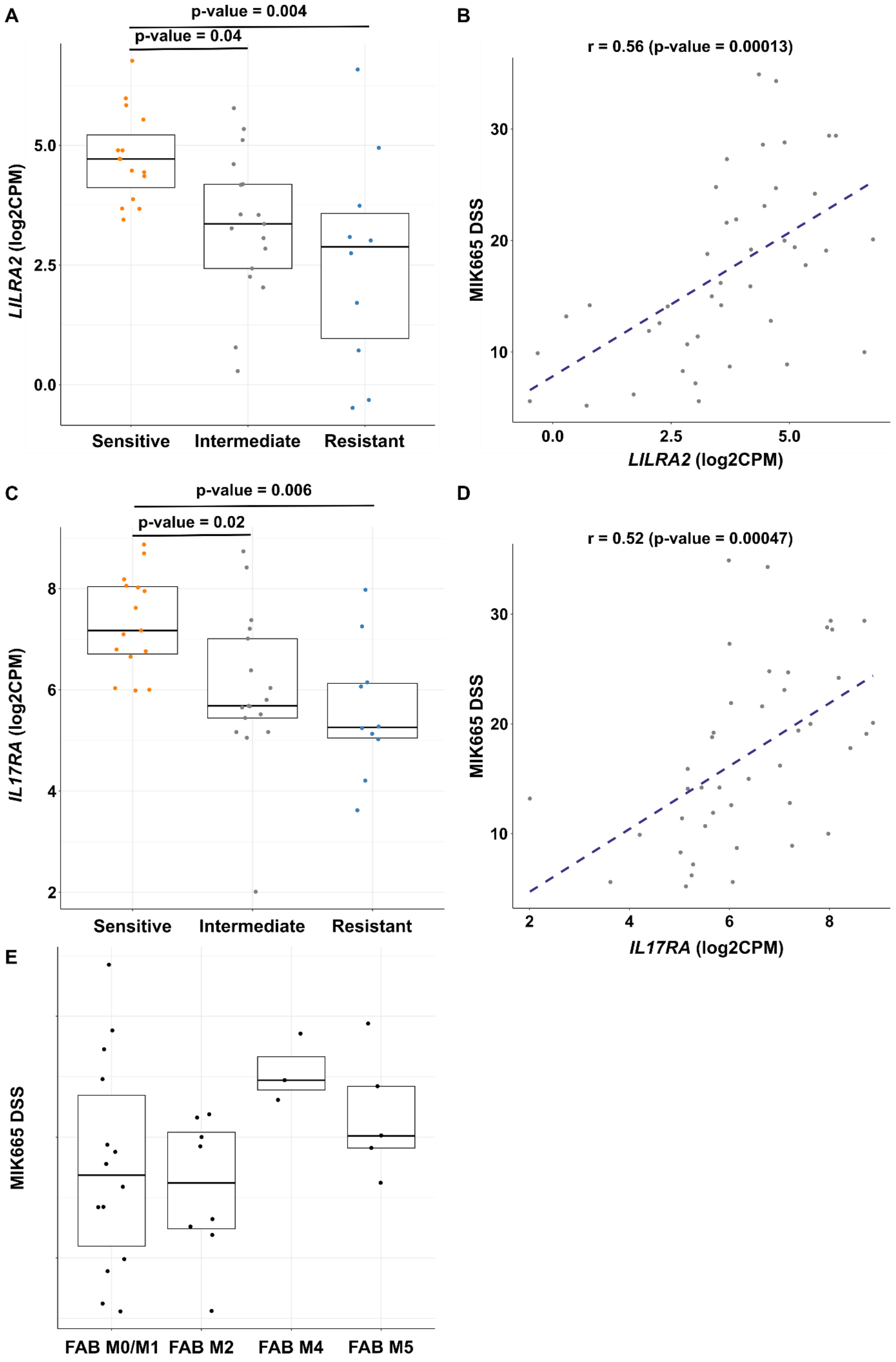
A
p-value = 0.004
p-value = 0.04
LILRA2 (log2CPM)
5.0
2.5
0.0
Sensitive
Intermediate
Resistant
B
r = 0.56 (p-value = 0.00013)
MIK665 DSS
30
20
10
0.0
2.5
5.0
LILRA2 (log2CPM)
C
p-value = 0.006
p-value = 0.02
IL17RA (log2CPM)
8
6
4
2
Sensitive
Intermediate
Resistant
D
r = 0.52 (p-value = 0.00047)
MIK665 DSS
30
20
10
2
4
6
8
IL17RA (log2CPM)
E
MIK665 DSS
FAB M0/M1
FAB M2
FAB M4
FAB M5

**Supplementary Figure 9. Differentiation-associated genes correlate with MIK665 sensitivity. A)** The median level of expression of *LILRA2* decreases significantly with MIK665 resistance (anova p-value = 0.0035), **B)** and is strongly correlated with response to MIK665 (r = 0.56, p-value = 0.00013). **C)** The median level of expression of *IL17RA* decreases significantly with MIK665 resistance (anova p-value = 0.0036), **D)** and is strongly correlated with response to MIK665 (r = 0.52, p-value = 0.00047). **E)** AML samples with a more differentiated FAB type (M4/M5) have higher MIK665 DSS values than samples with a less differentiated FAB type (M0/M1/M2). Correlation tests were performed using the Spearman method, and pairwise comparisons using the Tukey test.

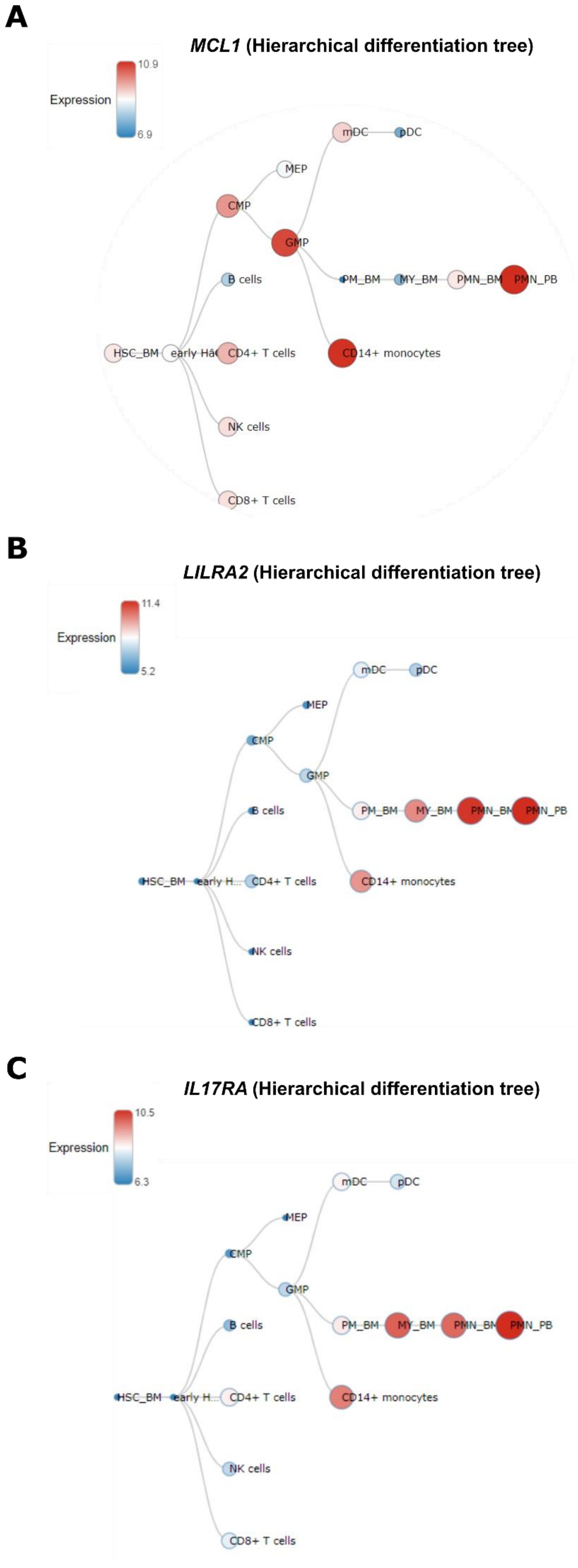
A
MCL1 (Hierarchical differentiation tree)
Expression
10.9
6.9
mDC
pDC
MEP
CMP
GMP
B cells
PM_BM
MY_BM
PMN_BM
PMN_PB
HSC_BM
CD4+ T cells
CD14+ monocytes
NK cells
CD8+ T cells
B
LILRA2 (Hierarchical differentiation tree)
Expression
11.4
5.2
mDC
pDC
MEP
CMP
GMP
B cells
PM_BM
MY_BM
PMN_PB
HSC_BM
early H...
CD4+ T cells
CD14+ monocytes
NK cells
CD8+ T cells
C
IL17RA (Hierarchical differentiation tree)
Expression
10.5
6.3
mDC
pDC
MEP
CMP
GMP
B cells
PM_BM
MY_BM
PMN_PB
HSC_BM
early H...
CD4+ T cells
CD14+ monocytes
NK cells
CD8+ T cells

**Supplementary Figure 10. Hematopoietic trees demonstrating the increased expression of A)** *MCL1*, **B)** *LILRA2* and **C)** *IL17RA* with differentiation in normal human hematopoiesis, notably towards cells of the monocytic and polymorphonuclear lineages (Source: HemaExplorer on BloodSpot).

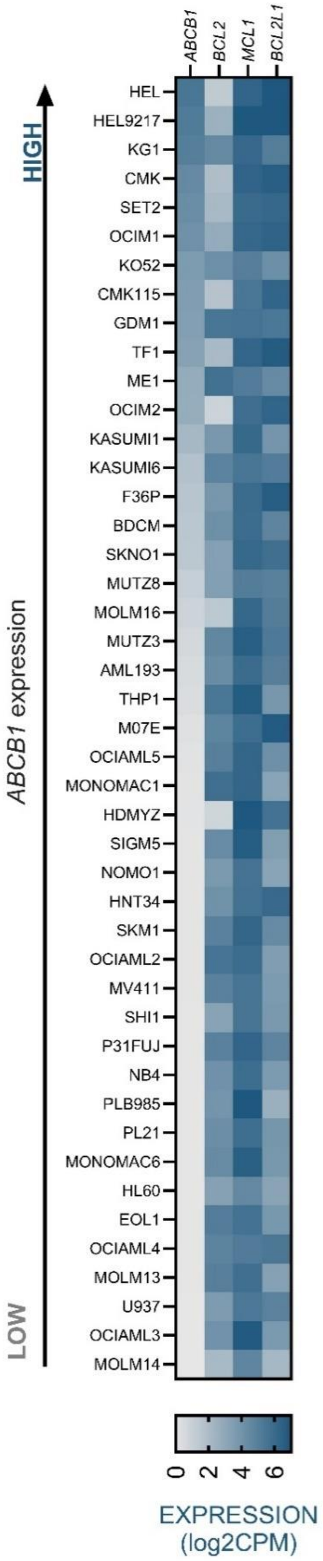


**Supplementary Figure 11. Heatmap showing *ABCB1*, *BCL2*, *MCL1* and *BCL2L1* expression in 45 AML cell lines.** The cell lines are sorted by *ABCB1* expression in log2CPM (Source: DepMap).

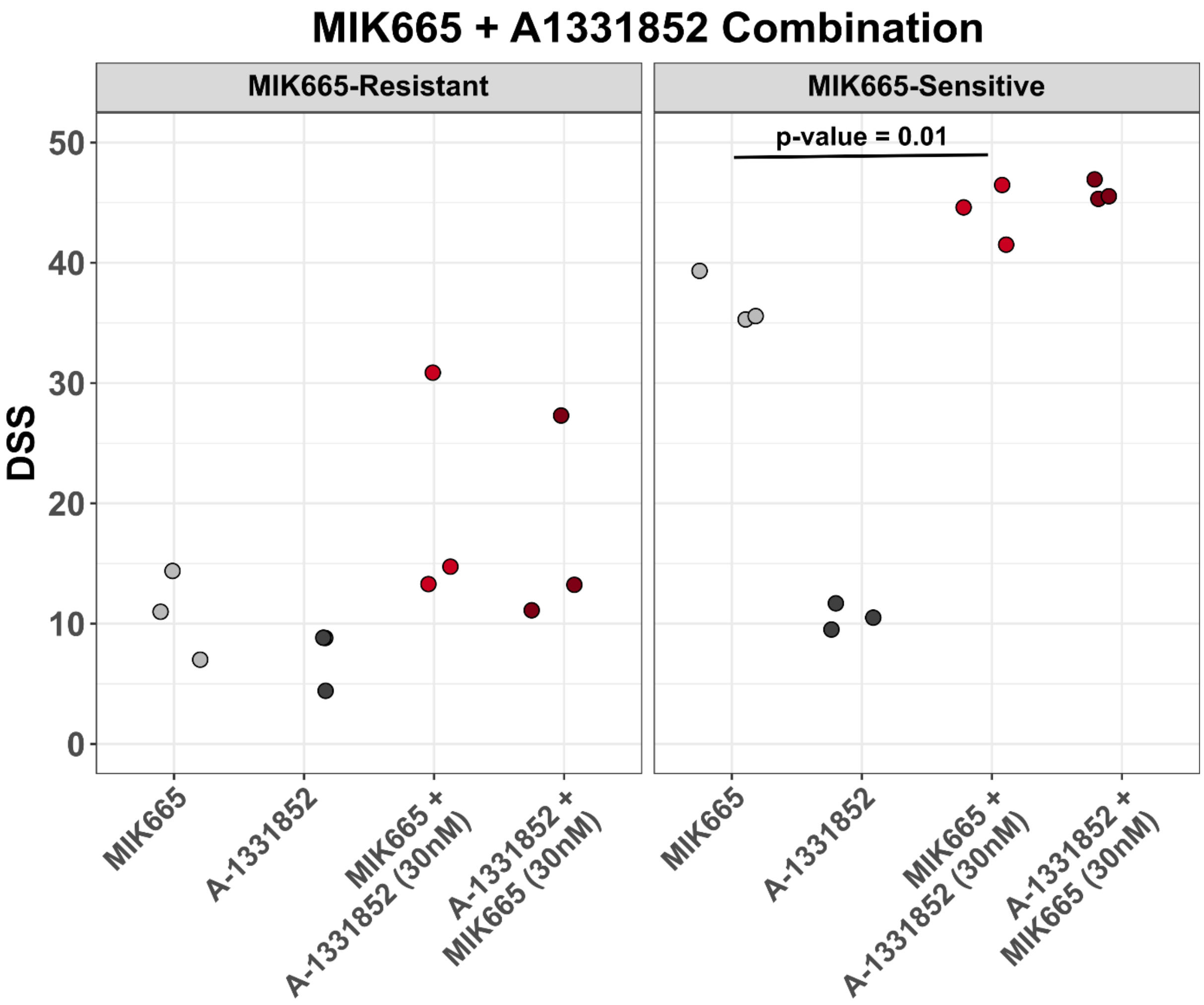


**Supplementary Figure 12. Dot plot showing the DSS of MIK665 and BCL-XL inhibitor A1331852 alone or in combination in AML samples with MIK665 resistance and MIK665 sensitivity.** MIK665 resistance was defined as high *ABCB1*: log2CPM > 6, and MIK665 sensitivity was defined as low *ABCB1*: log2CPM < 6. Grey dots represent the DSS values of the single agents, whereas red dots represent the DSS values of the combination (where one drug is increased along its concentration range while the other drug is fixed at 30 nM). Significance was evaluated using the paired sample t-test.

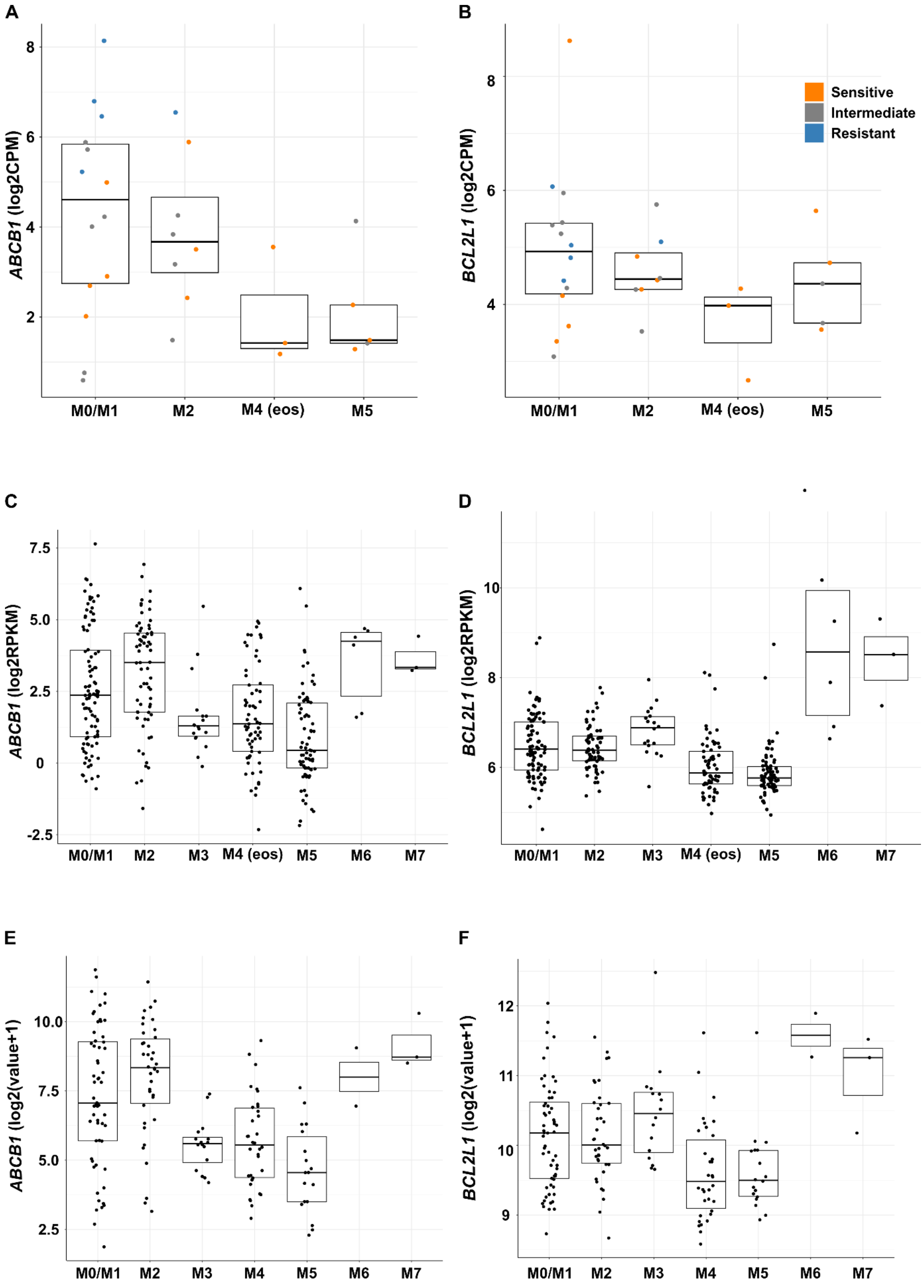


**Supplementary Figure 13. Expression of *ABCB1* and *BCL2L1* across the FAB types of primary AML samples.** *ABCB1* and *BCL2L1* show higher levels of expression in M0/M1 and M2 samples compared to M4 and M5 samples in the **A-B)** FIMM, **C-D)** BEAT, and **E-F)** TCGA cohorts. FIMM samples are colored according to their response levels to MIK665. Significance was evaluated using the one-way anova test, followed by the Tukey test for pairwise comparisons.

### Supplementary Materials and Methods

#### Antibody staining and flow cytometry-based readouts

Following drug treatments, sample plates were centrifuged for 6 minutes at 500g and the supernatant removed by flipping the plate. Cells were then stained in staining buffer (5% FBS in PBS) containing CD45 and CD14 antibodies (BD Biosciences, San Diego, CA, USA), and incubated for 30 minutes at room temperature in the dark (Supplementary Table 3). Each well was stained with 25μL of the antibody mix.

After staining, the cells were centrifuged at 500 g and the supernatant removed. This was followed by a viability-staining step for 10 minutes in room temperature in the dark with Annexin-V and 7-AAD to identify apoptotic and dead cells, respectively. Each well was stained with 25 μL of the viability mix added to the 1x Annexin V Binding Buffer diluted with MQ water (Supplementary Table 3).

After the viability-staining step, the plates were screened in the iQue Screener PLUS (Intellicyt) instrument. The sipping parameters per well were set to 17 s with a pump speed of 29 rpm. Once a plate was read, cell gating was performed using the in-built ForeCyt software (Intellicyt). An illustration of the gating strategy is available in Supplementary Figure 2.

#### Differential gene expression analysis

Prior to differential gene expression analysis, non-protein coding genes were filtered out. Afterwards, trimmed mean of M-values (TMM) normalization was performed using the edgeR package in R and counts per million (CPM) values were computed (24). To filter out genes with low expression across the samples, only those with a CPM value larger than 1 in at least 50% of the samples in the analysis were retained. A limmavoom pipeline was used for log2CPM calculation and regression modeling and differentially expressed genes were defined as those having an FDR < 0.1.

##### Western blotting

Cells were lysed using RIPA buffer supplemented with phosphatase inhibitor and phenylmethylsulphonyl fluoride (Cell Signaling Technology, Danvers, MA). Proteins were quantified using the Pierce BCA protein assay kit (Thermo-Fisher, Waltham, MA), and subsequently separated by SDS-PAGE and transferred to nitrocellulose membranes. Signals were acquired on an Odyssey scanner and blots were quantified using the Image Studio Lite software (LI-COR Biosciences, Lincoln, NE). The antibodies used are reported in Supplementary Table 4.

##### Quantitative reverse transcription polymerase chain reaction (RT-qPCR)

RT-qPCR was performed with reference to MIQE guidelines (1). RNA was extracted from MNCs using Macherey-Nagel NucleoSpin RNA kit (Düren, Germany), quantified on a Qubit fluorometer (ThermoFisher), and the quality assessed using an Agilent Bioanalyzer (Santa Clara, CA). cDNA was prepared using gene specific primers (Supplementary Table 6) and SOLIScript reverse transcriptase (Solis Biodyne, Tartu, Estonia). RT-qPCR was performed with HOT FIREPol EvaGreen qPCR Mix Plus (Solis Biodyne), using four reference genes: SH3D19, HNRNPC, EIF4B and NONO, with SH3D19 being excluded due to low expression. Reference genes were found to have low M-values in CFX Maestro software (Bio Rad, Hercules, CA). Expression values per sample were normalized to the sample with the lowest expression score, and the resulting fold change values were log transformed.

##### Knockout of *ABCB1* in HEL cells using CRISPR/Cas9

Plasmids were purified using NucleoSpin Plasmid mini kit (Machery-Nagel, Düren, Germany) and the yield of each plasmid was determined using Qubit fluorometer (ThermoFisher). HEK293-FT cells were seeded in 100x20 mm plates at a concentration of 0.5 M/ml in a total of 10 ml of DMEM. After 24 h, cells were transfected with 10 µg lentiCRISPRv2GFP-ABCB1, 6 µg psPAX2, and 4 µg pCMV-VSV-G using a calcium phosphate transfection kit (Promega) with the addition of 600 µM of chloroquine. The mixture was added dropwise while swirling to homogenize. Transfections with lentiCRISPRv2GFP-NTC or LEGO-v2 plasmids were used as controls. 6 h after transfection, media was replaced. Virus was harvested after 48 h incubation at 37 $^{o}$C and filtered using a 0.45 µm filters.

HEL cells were seeded in a 24-well plate at 0.2 M/ml in 100 µL of RPMI. Viral suspension was added at a ratio of 4:1. Cells were kept in culture, and when necessary, were split 1:3 with PBS wash. 1M HEL cells were taken for fluorescence-activated cell sorting using a BD Influx cell sorter (Franklin Lakes, NJ, USA), and GFP-positive cells were seeded in 96-well V-bottom plates (ThermoFisher) at a density of one cell per well. Cells were maintained in 100 µL of complete RPMI media per well (50 µL of fresh media and 50 µL of conditioned media from the parental cells) and expanded when needed. PCR was used to identify *ABCB1* knockout which was confirmed using sanger sequencing and western blotting.
